# segcsvd_PVS_: A convolutional neural network-based tool for quantification of enlarged perivascular spaces (PVS) on T1-weighted images

**DOI:** 10.1101/2025.07.29.25332360

**Authors:** Erin Gibson, Joel Ramirez, Lauren Abby Woods, Stephanie Berberian, Julie Ottoy, Christopher J.M. Scott, Vanessa Yhap, Fuqiang Gao, Roberto Duarte Coello, Maria Valdes-Hernandez, Anthony E. Lang, Carmela M. Tartaglia, Sanjeev Kumar, Malcolm A. Binns, Robert Bartha, Sean Symons, Richard H. Swartz, Mario Masellis, Navneet Singh, Bradley J. MacIntosh, Joanna M. Wardlaw, Sandra E. Black, ONDRI Investigators, ADNI, CAIN Investigators, colleagues from the Foundation Leducq Transatlantic Network of Excellence, Andrew SP Lim, Maged Goubran

## Abstract

**Introduction:** Enlarged perivascular spaces (PVS) are imaging markers of cerebral small vessel disease (CSVD) that are associated with age, disease phenotypes, and overall health. Quantification of PVS is challenging but necessary to expand an understanding of their role in cerebrovascular pathology. Accurate and automated segmentation of PVS on T1-weighted images would be valuable given the widespread use of T1-weighted imaging protocols in multisite clinical and research datasets.

**Methods:** We introduce segcsvd_PVS_, a convolutional neural network (CNN)-based tool for automated PVS segmentation on T1-weighted images. segcsvd_PVS_ was developed using a novel hierarchical approach that builds on existing tools and incorporates robust training strategies to enhance the accuracy and consistency of PVS segmentation. Performance was evaluated using a comprehensive evaluation strategy that included comparison to existing benchmark methods, ablation-based validation, accuracy validation against manual ground truth annotations, correlation with age-related PVS burden as a biological benchmark, and extensive robustness testing.

**Results:** segcsvd_PVS_ achieved strong object-level performance for basal ganglia PVS (DSC = 0.78), exhibiting both high sensitivity (SNS = 0.80) and precision (PRC = 0.78). Although voxel-level precision was lower (PRC = 0.57), manual correction improved this by only ~3%, indicating that the additional voxels reflected primary boundary- or extent-related differences rather than correctable false positive error. For non-basal ganglia PVS, segcsvd_PVS_ outperformed benchmark methods, exhibiting higher voxel-level performance across several metrics (DSC = 0.60, SNS = 0.67, PRC = 0.57, NSD = 0.77), despite overall lower performance relative to basal ganglia PVS. Additionally, the association between age and segmentation-derived measures of PVS burden were consistently stronger and more reliable for segcsvd_PVS_ compared to benchmark methods across three cohorts (test6, ADNI, CAHHM), providing further evidence of the accuracy and consistency of its segmentation output.

**Conclusions:** segcsvd_PVS_ demonstrates robust performance across diverse imaging conditions and improved sensitivity to biologically meaningful associations, supporting its utility as a T1-based PVS segmentation tool.

## Introduction

Enlarged perivascular spaces (PVS), also known as Virchow-Robin spaces, are small, fluid-filled structures that are commonly observed on brain magnetic resonance imaging (MRI), particularly in older adults and individuals with neurovascular and neurodegenerative diseases (Wardlaw et al., 2013). They are recognized as an important imaging marker of cerebral small vessel disease (CSVD), where their presence and burden are increasingly associated with poor sleep health, disease phenotypes, hypertension, age, and overall health indicators (Waymont et al., 2024). Accurate detection and quantification of PVS are crucial for diagnosis, monitoring disease progression, and research into the underlying mechanisms of these conditions (Wardlaw et al., 2019), necessitating the development of automated, reliable, and efficient tools for PVS segmentation.

PVS appear as small hypointense (dark) structures on T1-weighted images and hyperintense (bright) structures on T2-weighted images. While T2-weighted images are more sensitive to PVS, especially smaller PVS that may represent the earliest stages of pathology, T1-weighted images are more widely available due to their routine inclusion in structural imaging protocols. This broader accessibility creates a need for tools that segment PVS on T1-weighted images; however, this presents additional challenges, primarily due to their limited contrast, which hinders the accurate identification and delineation of PVS boundaries.

Existing methods for PVS segmentation typically rely on either filtering or deep learning approaches, each having distinct advantages and limitations (Waymont et al., 2024). Filtering approaches based on the Frangi (Frangi et al., 1998) or RORPO (Ranking Orientation Responses of Path Openings) filter (Merveille et al., 2017) rely on geometric properties such as tubular shape or “vesselness” to detect PVS. While these methods can perform well for single-site, artifact-free data, they often require manual parameter tuning to adapt to variations in image contrast or quality, which can limit their generalizability across datasets. Moreover, they are not specific to PVS and instead detect all tubular structures in the image, typically requiring additional false positive minimization (FPM) strategies. As such, there remains a need for more automated tools that approximate the behavior of these filters while improving consistency and scalability across large, heterogeneous datasets

Deep learning models, and convolutional neural networks (CNNs) in particular, have demonstrated success in medical imaging segmentation tasks (e.g., Gibson et. al., 2024; Mojiri et al., 2022; Ntiri et al., 2021). Although these models excel at learning complex features and are fully automated, their generalizability is often limited, particularly for datasets with imaging parameters or patient populations that differ from the training distribution. Additionally, generating the ground truth segmentations necessary for training these models is challenging due to the small size of PVS, which typically ranges between 2 and 5 mm in diameter. Manual approaches are time-consuming and prone to both inter- and intra-rater variability and error (Pham et al., 2022), where the limitations of human annotation prevent the reliable identification and delineation of subtle PVS structures. Semi-automated segmentation methods are more efficient (e.g., Ballerini et al., 2018) but are inherently biased by the underlying automated tool. Consequently, both manual and semi-automated approaches introduce variability and bias into the ground truth segmentations, presenting challenges for the objective training and validation of fully automated PVS segmentation tools.

While several fully automated deep-learning PVS segmentation tools have been described in the literature, currently only two are publicly available for T1-weighted images. SHIVA (Boutinaud et al. 2021), based on an autoencoder and U-net was trained exclusively on young, healthy subjects, limiting its applicability to broader populations. PINGU (Sinclair et al., 2024), based on the nnUNet framework (Isensee et al., 2021), was trained on a more diverse set of images spanning a wider range of ages, patient populations and imaging parameters. Both tools relied on manual segmentations for model training, with PINGU outperforming SHIVA in segmentation accuracy on their test sets. However, performance scores for PINGU remained relatively modest overall, with Dice scores ranging between approximately 0.3 and 0.5 on external validation datasets (Sinclair et al., 2024).

The current study aims to address ongoing challenges in PVS quantification through the development of segcsvd_PVS_ (segcsvd), a fully automated convolutional neural network (CNN)-based segmentation method for T1-weighted images. The segcsvd segmentation model builds on existing tools and incorporates robust training strategies to improve the accuracy and consistency of PVS segmentation. Performance is evaluated using both semi-automated and manual ground truth annotations, and benchmarked against existing methods, with a focus on: 1) segmentation accuracy, 2) the strength of detected biologically relevant associations (i.e., age-related changes in PVS measures), and 3) robustness to variations in image contrast and noise.

This study makes three key contributions. First, it integrates anatomical priors with advanced data augmentation strategies and leverages a large, diverse multisite dataset (n = 168) to support model generalizability. Second, it utilizes an extensively refined semi-automated ground truth dataset, designed to provide a consistent training signal while minimizing label noise. Third, it introduces a comprehensive evaluation framework that extends beyond conventional performance metrics to assess both the stability of segmentation output and its correspondence with biologically meaningful variables. To our knowledge, such validation has not been undertaken in prior work, leaving important aspects of robustness and biological relevance underexplored in this context.

## Methods

### MRI data

#### Training, validation and test datasets

A large dataset of T1-weighted MRI scans acquired at 3T (*n* = 1670) was assembled from five diverse, multisite studies (Table 1). These studies spanned a wide range of patient populations, including individuals with sleep apnea, cardiovascular risk factors, carotid stenosis, cerebrovascular disease, Parkinson’s disease and healthy controls (mean age = 63.2 ± 9.4).

**Table 1.** Key imaging characteristics for the T1-weighted images used for model development and validation. Bolded n-values indicate data used exclusively for model validation. Abbreviations: (LD) Foundation Leducq Transatlantic Network of Excellence on the Role of the Perivascular Space in Cerebral Small Vessel Disease; (CAHHM) Canadian Alliance for Healthy Hearts and Minds; (CAIN) Canadian Atherosclerosis Imaging Network Project 1; (ADNI) Alzheimer’s Disease Neuroimaging Initiative 3; (ONDRI) Ontario Neurodegenerative Disease Research Initiative; (SBH) Sunnybrook Health Sciences Centre; (TWH) Toronto Western Health; (CR_GM-sCSF_) contrast ratio for GM relative to sCSF; (CR_GM-sCSF_) contrast ratio for WM relative to sCSF.

| Dataset | RORPO<sub>GT</sub> | Manual<sub>GT</sub> | Patient Population / Datasets | Voxel Dimensions (mm) | CR<sub>GM→CSF</sub> | CR<sub>WM→CSF</sub> | %vCSF | n |
| --- | --- | --- | --- | --- | --- | --- | --- | --- |
| **LD**<br>single site | yes | no | sleep apnea | 1.0 x 1.0 x 1.0 | -0.97 ± 0.07 | -1.75 ± 0.06 | 2.7 ± 1.4 | 108 |
| **CAHHM**<br>multiple sites | yes | no | individuals with varying cardiovascular risk factors | 0.9 x 0.9 x 0.9 | -0.84 ± 0.07 | -1.83 ± 0.06 | 2.1 ± 0.8 | 30 |
|  | no | no |  |  | -0.94 ± 0.07 | -1.75 ± 0.08 | 1.8 ± 0.8 | **764** |
| **CAIN**<br>multiple sites | yes | no | non-surgical carotid stenosis | 1.0 x 1.0 x 1.0 | -0.98 ± 0.06 | -1.78 ± 0.08 | 4.0 ± 1.8 | 34 |
| **ONDRI**<br>two sites | yes | no | cerebrovascular disease, Parkinson's disease | 1.0 x 1.0 x 1.0 | -0.91 ± 0.08 | -1.86 ± 0.08 | 3.2 ± 1.6 | 34 |
| **ADNI**<br>multiple sites | no | no | cognitively normal (ADNI screening scan) | 1.0 x 1.0 x 1.0 | -0.97 ± 0.06 | -1.81 ± 0.08 | 2.4 ± 1.1 | **350** |
| **test**<br>multiple sites | yes | no | LD ($n = 4$),<br>CAHHM ($n = 2$),<br>CAIN ($n = 1$),<br>ONDRI site 1 ($n = 10$),<br>ONDRI site 2 ($n = 14$) | 0.9 x 0.9 x 0.9 ($n = 2$),<br>1.0 x 1.0 x 1.0 ($n = 28$) | -0.93 ± 0.10 | -1.85 ± 0.09 | 3.2 ± 1.6 | **31** |
| **test<sub>6</sub>**<br>multiple sites | yes | yes | 6 slices from the test dataset |  |  |  |  |  |

This dataset was characterized by variability in both anatomical and imaging characteristics, providing a robust foundation for the training and validation of segcsvd. Anatomical variability was quantified using ventricular cerebrospinal fluid (vCSF) volumes, calculated as a percentage of the total intracranial volume, which indexed widespread neurodegenerative anatomical changes.

Given the large number of contributing sites to this dataset, each with distinct acquisition protocols, conventional reporting of individual T1-weighted imaging parameters (e.g., TR, TE, TI, flip angle) was not feasible or informative. Instead, variability in the T1-weighted imaging properties was quantified by calculating contrast ratios (CR) on masked, standardized (z-scored) T1-weighted images. These CRs were calculated for both gray matter (GM) and white matter (WM) relative to sulcal cerebrospinal fluid (sCSF), and were defined as:

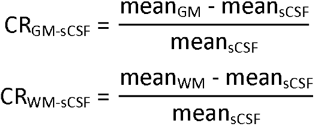

Together, these contrast ratios provided an index of the relative differentiation of hypointense (i.e. sCSF) voxels from surrounding WM and GM tissue, a differentiation critical for this PVS segmentation task, which involves segmentation of hypointense structures from surrounding voxels.

T1-weighted images acquired with non-isotropic voxel resolution were resampled to have isotropic voxel dimensions of either 0.9 mm or 1.0 mm, based on whichever was closest to the original in-plane acquisition resolution. This standardization was performed to facilitate downstream analyses of PVS morphological features while also ensuring a multi-resolution training framework designed to promote generalization across T1-weighted images acquired with approximately 1 mm voxels, as is typical in clinical and research settings.

Semi-automated ground truth PVS segmentations (*n* = 206; RORPO_GT_), described below, were generated for four of the datasets and were used for both model training and ablation-based validation of segcsvd. This set was randomly divided into training (*n* = 168), validation (*n* = 7), and test datasets (*n* = 31). The small validation dataset was used only to establish coarse limits for the number of training epochs, while the test dataset was reserved exclusively for the ablation-based validation. The test dataset included 10 scans from a hold-out site within the ONDRI dataset that were not used for model training. Imaging characteristics, including contrast ratios and percent vCSF, showed no notable differences between the hold-out and included sites and therefore the data were consolidated into a single “test” dataset.

Manual ground truth PVS segmentations (*n* = 31 × 6 slices; tracer_1_ and tracer_2_), described below, were generated by two human tracers for six slices in the test dataset. This subset, referred to as the “test_6_” dataset, was used to evaluate the accuracy of segcsvd relative to human annotators.

Two large datasets (CAHHM: n = 764; ADNI: *n* = 350) without ground truth data were also included. These datasets were used to evaluate the stability of both the voxel-level segmentation and age-related associations with PVS burden. The ADNI dataset was not used for model training and thus served as a large independent test dataset in this context.

In addition to the T1-weighted images, binary WMH segmentations masks derived from FLAIR images coregistered to the T1 image were generated for each dataset, following previously described methods (Gibson et al., 2024).

### Semi-automated ground truth PVS segmentations (RORPO_GT_)

A high-quality ground truth dataset (*n* = 206) was generated using a semi-automated procedure based on the RORPO (Merveille et al., 2017) filter (Figure 1). The RORPO filter was selected because previous work has shown that it outperforms other filters including the Frangi filter for images with isotropic voxels (Duarte Coello et al., 2024).

**Figure 1.**
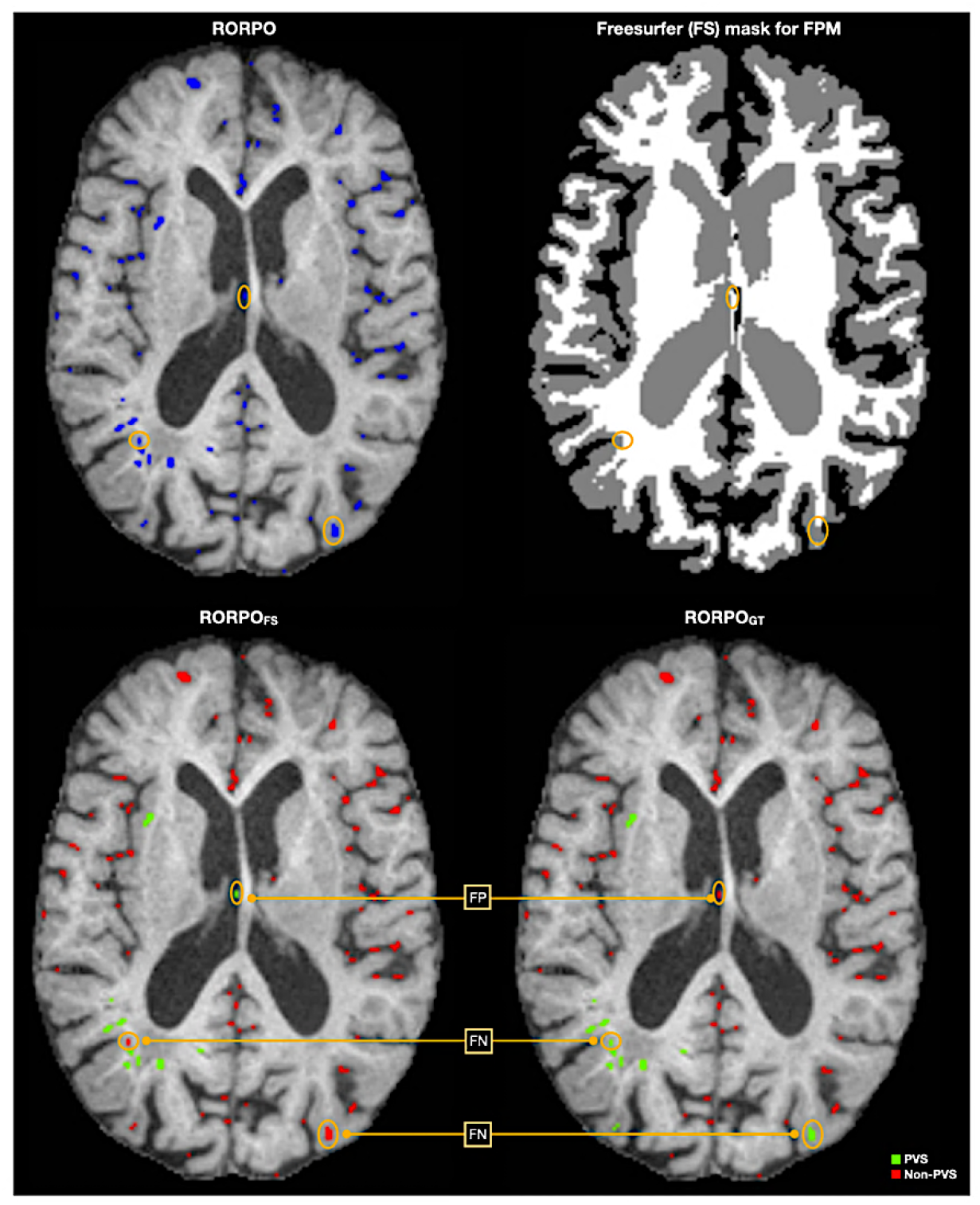
Semi-automated process to generate ground truth segmentations for model training and initial validation. The RORPO filter was first applied to extract all tubular structures from the T1-weighted images (blue voxels). The FreeSurfer (FS) “aseg” mask was then as a false positive minimization strategy, identifying PVS-prone regions and relabelling all RORPO-detected voxels as either probable PVS (green) or non-PVS (red) voxels (RORPO_FS_). This eliminated most false positives (FP) from the RORPO output but also introduced some false negatives (FN). The RORPO_FS_ segmentation was then manually edited, relabelling these remaining false negatives and false positives to create the final semi-automated ground truth segmentations (RORPO_GT_).

The semi-automated procedure consisted of several steps. First, FreeSurfer’s SynthSeg tool (Billot et al., 2023) was applied to the T1 image and the resulting segmentation was used to mask the T1 image, removing non-brain voxels. Anisotropic diffusion filtering (Perona and Malik, 1990) was then applied to the masked T1 image to reduce noise while preserving the edges of PVS structures. The RORPO filter was subsequently applied to the masked, diffusion-filtered T1 image.

Specification of several parameters was required for both the anisotropic diffusion and RORPO filter, and these were defined manually for each dataset (Table 2). While this introduced a degree of subjectivity in the generation of the semi-automated ground truth segmentations, these segmentations served primarily as the training signal for the segcsvd model. Slight variations in these parameters across datasets were therefore intended to promote learning of more fundamental, invariant features that define PVS, thereby enhancing consistency in performance across datasets – an approach that has performed well in our previous work (Gibson et al., 2024).

**Table 2.** Anisotropic diffusion and RORPO filter parameters for each dataset. For the Anisotropic diffusion filter, a time step of 0.025 was used for all datasets. For the RORPO filter, a dilation size of 0, minimum scale of 1, and binary threshold of 18 were used for all datasets.

| dataset | Anisotropic Diffusion Filter |  | RORPO Filter |  |
| --- | --- | --- | --- | --- |
|  | conductance | number of iterations | factor | nbScales |
| LD | 1.25 | 25 | 2.0 | 4 |
| CAHHM | 1.00 | 25 | 1.7 | 5 |
| CAIN | 1.00 | 22 | 1.7 | 5 |
| ONDRI | 1.25 | 25 | 2.0 | 4 |
| ADNI | 1.00 | 22 | 1.7 | 5 |

Application of the RORPO filter identified all hypointense tubular structures within the filter’s parameters, which included true PVS as well as numerous false positives from non-PVS structures (Figure 1 “RORPO”). To minimize these false positives, the “aseg” mask from the full FreeSurfer pipeline (Fischl et al., 2002) was used as a false positive minimization strategy (FPM), where RORPO-identified objects not connected in 3D to a voxel within a PVS-relevant region (i.e. thalamus, hippocampus, white matter, and basal ganglia) were relabeled as non-PVS (Figure 1 “RORPO_FS_”). Additionally, any voxels identified as WMH in the binary WMH segmentation mask were relabeled as non-PVS.

While this false positive minimization step was largely effective, it nevertheless introduced some errors, either by erroneously removing true PVS voxels or by failing to exclude non-PVS voxels. These errors were corrected through manual editing by a trained image analyst (L.A.W.). Manual editing was performed in ITK-SNAP for all images (n = 207) following a standardized protocol to ensure consistent application of inclusion and exclusion criteria. Tri-planar views (axial, sagittal, coronal) were used to assess each voxel in anatomical context. Intensity windowing was adjusted relative to fixed CSF and white matter peaks in the T1-weighted image histogram to provide consistent contrast and PVS visibility. Editing was restricted to the removal of false positive PVS objects and inclusion of false negative non-PVS objects based on anatomical plausibility and expert visual assessment in the context of surrounding structures. The boundaries of RORPO-detected structures were not modified (Figure 1, “RORPO_GT_”). This decision reflected practical constraints due to the time-intensive nature of manual refinement, as well as the difficulty of achieving consistent manual boundary delineation for small structures like PVS, where manual adjustments can introduce more variability than they resolve.

The FreeSurfer aseg (FS) mask was used exclusively for generating the semi-automated RORPO_GT_ segmentations, as the full FreeSurfer pipeline is time-intensive, requiring approximately 14 hours per subject to complete. In all other cases, the FreeSurfer SynthSeg (FSss) mask (Billot et al., 2023), which can be generated in approximately 2 minutes, was used as a more efficient and practical alternative (i.e., for RORPO_FSss_/RORPO_SEED_, described below).

### Manual ground truth segmentations

#### tracer1 and tracer2

Generating manual ground truth segmentations is both time consuming and prone to low inter-rater reliability (Pham et al., 2022). As a result, two trained analysts (L.A.W. and S.B.) produced manual segmentations for limited number of slices (*n* = 6) per subject in the test dataset (*n* = 31), creating the test_6_ dataset.

The six slices were pseudo-randomly selected, with three consecutive axial slices chosen at the level of the basal ganglia and an additional three consecutive axial slices randomly selected within a 20-slice range above or below the initial set. This approach ensured a focus on the basal ganglia, a region of clinical significance for PVS, while also incorporating more challenging regions outside this area, including the centrum semiovale and deep white matter areas. By limiting the selection to a smaller subset of slices, this approach balanced the need for detailed, high-quality annotations with practical constraints such as time and tracer fatigue.

Manual tracing was performed in ITK-SNAP using a standardized procedure to ensure consistent application of inclusion and exclusion criteria. Tri-planar views (axial, sagittal, coronal) were used to assess each voxel in anatomical context. Intensity windowing was adjusted relative to fixed CSF and white matter peaks in the T1-weighted image histogram to provide consistent contrast and PVS visibility. To support accurate and systematic manual segmentation, a seed image (RORPO_SEED_) was generated, consisting of single-voxel centroids from the RORPO segmentation and sulcal CSF voxels derived from the FreeSurfer SynthSeg mask (Figure 2). This seed image served as a reference to assist tracers by highlighting candidate PVS objects as well as potential false positives originating from sulcal CSF. The use of single-voxel centroids, rather than full RORPO segmentations, was a deliberate choice to avoid biasing the shape or extent of the manually traced PVS, while ensuring that full manual tracing was still required. While this approach may introduce some risk of confirmation bias toward RORPO-detected structures, this was mitigated by explicit instructions to review all voxels in each slice, to trace all visible PVS (including those not associated with a centroid), and to exclude any non-PVS voxels associated with a centroid. This strategy was guided by both methodological and practical considerations, leveraging an established PVS quantification method to enhance manual segmentation accuracy, while still requiring full manual delineation of PVS boundaries and extent.

**Figure 2.**
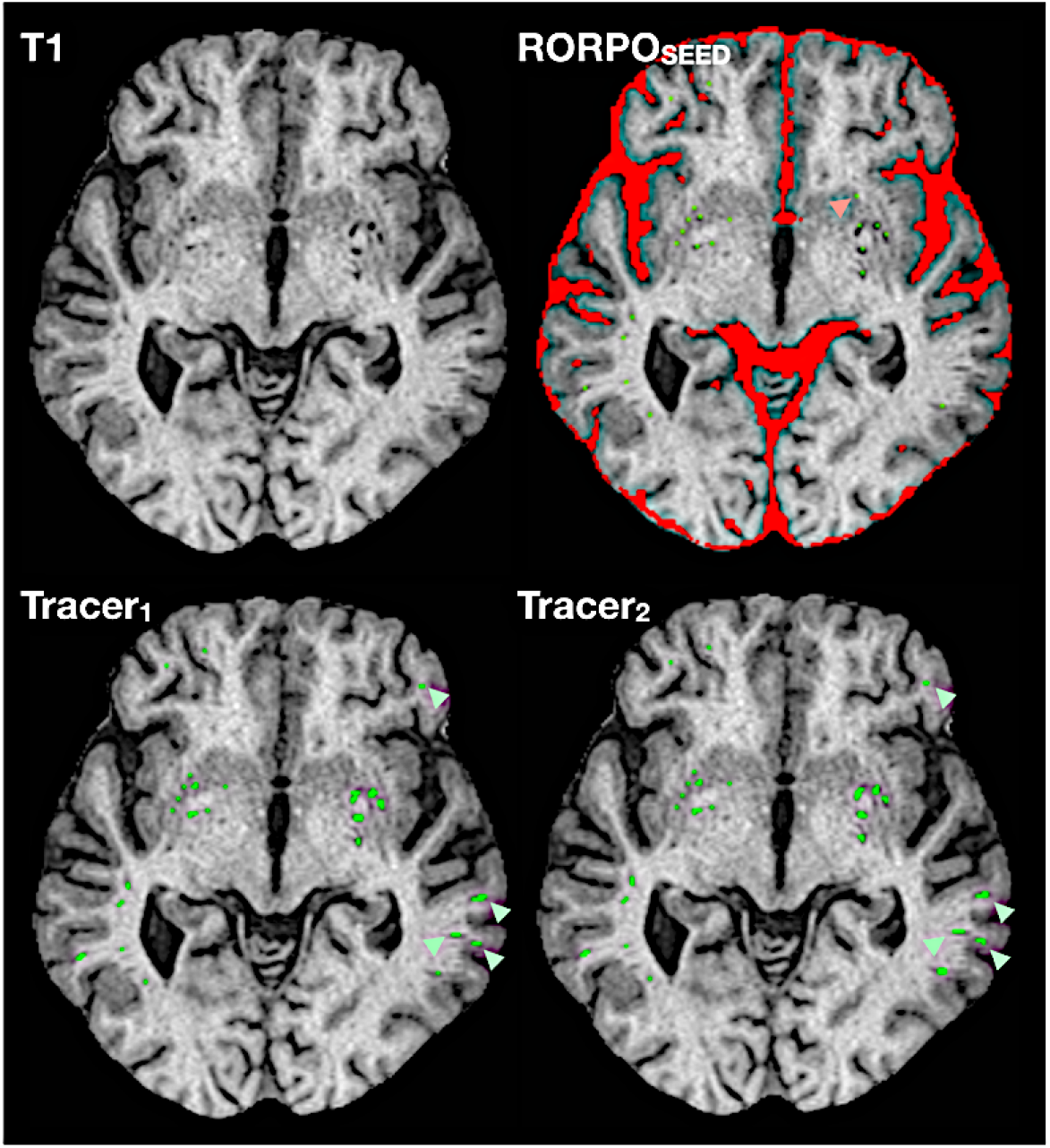
Manual segmentations for the test_6_ dataset. Single-voxel centroids from the RORPO segmentation, along with sulcal CSF from the FreeSurfer SynthSeg (FSss) mask, were combined to create an initial “RORPO_SEED_” image. This provided a reference for the manual segmentations, enabling the tracers to leverage PVS detection information from the RORPO filter, without biasing their precise delineation of individual PVS. With the RORPO_SEED_ image as a reference, the tracers manually traced PVS on each slice, including PVS without RORPO-centroids (light green arrows), and excluding non-PVS with RORPO centroids (light red arrow).

#### Consensus manual ground truth segmentation (consensus_GT_)

A consensus manual ground truth segmentation was created that included the union of all voxels identified by tracer_1_ and tracer_2_. While the term “consensus” implies agreement between tracers that is often operationalized as the intersection of labeled voxels, in the present work, the union of all voxels labeled by either tracer was used, prioritizing sensitivity to all manually segmented PVS voxels. In this context, the union can be interpreted as a more inclusive consensus, based on the rationale that any voxel identified by either tracer is likely to represent true PVS, given the known challenges of manually detecting small, subtle structures (Pham et. al., 2022).

### Model design and training

#### Model inputs

The segcsvd segmentation model is built upon a three-stage hierarchical framework that incorporates information from other segmentations to enhance PVS segmentation.

In the first stage, SynthSeg, a recently validated CNN-based tool (Billot et al., 2023) from the FreeSurfer software suite (Fischl, 2012) was utilized. This tool was developed using a large synthetic dataset and enables fast and robust segmentation of cortical and subcortical structures for diverse clinical scans of any contrast and resolution. segcsvd leverages the SynthSeg segmentation to generate a regional mask consisting of hippocampal and sulcal cerebrospinal (sCSF) voxels. This regional mask provides valuable spatial context to inform and constrain the subsequent PVS segmentation task, where the hippocampus acts as a key anatomical reference point and sCSF serves as a marker for the perimeter of cortical gray matter. These regions were selected because they provide relevant spatial context but are also sufficiently distanced from the white matter compartment to minimize the potential impact of any errors in the SynthSeg output on the PVS segmentation task.

In the second stage, a WMH segmentation mask is generated from a co-registered FLAIR image, using a previously validated method (Gibson et al., 2024). This step is optional and is only performed for populations with WMH to prevent the misclassification of WMH-related T1 hypointensities as PVS. For populations without WMH, an empty (all-zero) image can be used instead. This image is then integrated into the regional mask, using separate labels to denote sulcal CSF, hippocampal, and WMH voxels. Although this approach does not aim to segment PVS in WMH, the limited presence of PVS in WMH in the training data led to the decision to exclude these voxels from the ground truth segmentations for this work. If necessary, alternative methods or ad-hoc approaches, could address this limitation for future studies.

In the third stage, the regional mask and T1 image are used as dual inputs to a 3D CNN specifically optimized for full-brain PVS segmentation. Standard preprocessing, consisting of brain masking, bias correction, and intensity standardization, is performed to enhance consistency across datasets prior to inputting the T1-weighted images into the CNN. This three-stage approach explicitly combines relevant anatomical and WMH context with the contrast characteristics of the T1 image to improve the accuracy and sensitivity of the PVS segmentation.

#### Model architecture

The segcsvd segmentation model is similar to a model recently developed and validated for WMH segmentation (Gibson et al., 2024). Both are based on the U-Net architecture with residual units (Falk et al., 2019), which is a convolutional neural net (CNN) designed specifically for medical image segmentation tasks. It is composed of encoder and decoder pathways with skip connections to link the corresponding layers. These skip connections allow the network to capture both local and global features within the input data. The encoder pathway progressively downsamples the input data to extract hierarchical features at multiple spatial scales, while the decoder pathway upsamples the extracted features to generate the final segmentation output. This model, implemented in PyTorch using the MONAI toolkit (Cardoso et al., 2022), was configured using: 3 spatial dimensions; 2 input channels; a channel progression of 32, 62, 128, 256, 320; kernel size of 3, strides of 2, 2 residual units, batch normalization and a dropout rate of 0.1. The model takes as input both a masked and bias-corrected T1 image and a regional mask (described above)

#### Model training, ensembling, and prediction

Three separate models were trained using the Tversky loss function, which is an extension of the Dice coefficient that measures the similarity between two samples. The Tversky loss (TL) is defined as:

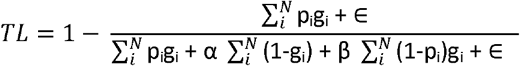

where:

p_i_ and g_i_ are the predicted and ground truth binary values at each pixel i

α and β are parameters that control the relative importance of false positives and false negatives respectively

∈ is a small constant added for numerical stability

N is the total number of pixels

When beta is equal to alpha, the Tversky loss function is equivalent to the Dice coefficient and provides a balanced weighting between false positives and false negatives. When beta is either greater than or less than alpha, more emphasis is placed on minimizing false negatives or false positives, respectively.

The three models were trained using different beta values for the Tversky loss function (β=0.50, 0.60, 0.65), and each model was initialized using a different random seed. This approach was designed to encourage each model to follow slightly different learning trajectories, thereby creating a diverse set of models, each characterized by subtle variations in their trade-off between precision and sensitivity. All other training hyperparameters remained the same across the three models. These hyperparameters included the use of the Adam optimizer with a learning rate of 0.00014, a training duration of 200 epochs, a patch size of 96 voxels^3^, a batch size of 4, a dropout rate of 0.1, and batch normalization. These parameters were selected empirically based on their effectiveness in preliminary experiments using a small subset of unedited RORPO_FS_ data (n < 40), conducted prior to generating the edited RORPO_GT_, as well as their demonstrated utility in related segmentation tasks (Gibson et al., 2024), with the rationale that parameters yielding favorable results in this context are likely to generalize well to the full-scale task. This strategy aimed to reduce the risk of overfitting, by selecting parameters with broad applicability rather than finely tuning parameters to a particular validation dataset.

Patch-based training was selected instead of whole-brain training, maintaining a consistent positive-to-negative label ratio for the PVS class as a strategy to address the issue of class imbalance. For each sample in the batch, four patches were extracted using a 3:1 positive-to-negative label ratio.

To enhance model generalization, several data augmentation transforms were randomly applied during training. These transforms included: contrast adjustment with gamma values randomly sampled between 0.6 and 1.2; the addition of Rician noise with a mean of 1.0 and standard deviation equal to 0.01% of the maximum voxel intensity of the image (noise factor = 0.0001); and random 90-degree rotations along each of three principal image axes. Image contrast augmentation was applied with of probability of 0.75, while all other augmentations were applied with a probability of 0.5. These transforms were selected to simulate the types of variability that are often present in clinical imaging datasets. Each image in the training dataset was sampled six times under these augmentations within each training epoch to ensure the model encountered a wide variety of transformations within every training epoch.

After training, a final, composite ensemble model was generated from the three base models. This was accomplished by establishing limits on the number of training epochs that would be considered for model ensembling, to prevent inclusion of models prone to overfitting or underfitting. The lower and upper limits were set to 150 and 200 epochs, respectively. These limits marked the initial performance plateau for the validation data, where the Dice coefficient and sensitivity scores first stabilized, and after which, began to exhibit only marginal improvements. Four models were then randomly selected from within these limits for each base model to form the final composite model (*n* = 12).

Predictions were then generated from each model in the ensemble using a test-time augmentation strategy consisting of generating predictions under both default and enhanced (γ =1.6) contrast. This approach produced 24 predictions per voxel, which were averaged to form a consensus segmentation image. This image was then thresholded at 0.35 to produce the final binary PVS segmentation image. This threshold was selected empirically to provide a balance between thresholds that tended to achieve the highest dice scores (~0.4) and thresholds that achieved the highest sensitivity scores (~0.3), on the validation data and in similar segmentation tasks (Gibson et al. 2024).

The use of two Tversky beta values favoring sensitivity (β = 0.60, 0.65), along with a test-time augmentation strategy that included predictions under enhanced contrast using a gamma value (γ =1.6) beyond what was used during training, was intended to increase the sensitivity of PVS segmentation. This strategy was implemented *a priori*, based on preliminary observations that the semi-automated RORPO segmentations, which served as ground truth for model training, tended to be conservative and occasionally missed true PVS voxels.

On an older generation Intel i7-11700 CPU, segcsvd completes PVS segmentation in approximately four minutes, inclusive of the time required for FreeSurfer’s SynthSeg.

### Comprehensive evaluation framework for segcsvd

#### 1. Quantitative assessment of manual editing and tracing efforts (semi-automated and manual GT datasets)

To assess the amount of manual editing and tracing performed during the construction of the ground truth datasets, standard measures of agreement, described below, were calculated. Agreement between RORPO_FS_ and RORPO_GT_ assessed the extent of manual editing performed to generate the semi-automated ground truth segmentations. Agreement between RORPO_SEED_ and each tracer assessed the extent of manual tracing performed to generate the manual segmentations. Agreement between the two tracers assessed the consistency of the manual segmentations used to derive the manual ground truth dataset (consensus_GT_).

#### 2. Validation of segcsvd model design (test dataset)

To validate the design of the segcsvd model, performance was assessed for the full model and several ablated variants using the test dataset. These variants excluded key components individually, including the WMH segmentation mask, test-time intensity augmentation, epoch ensembling, and Tversky loss ensembling, to evaluate the impact of each design choice on segmentation accuracy. Performance metrics were calculated relative to RORPO_GT_, which served as the training target for segcsvd. Using RORPO_GT_ as the reference enabled a systematic evaluation of model design and input modifications, without the added variability introduced if using manual ground truth segmentations as the reference.

#### 3. Validation of segmentation accuracy for segcsvd and two benchmark tools (test_6_ dataset)

To evaluate segmentation performance, segcsvd was compared to two existing benchmark methods using the test_6_ dataset. Performance was assessed relative to the consensus manual segmentation (consensus_GT_), providing external validation of segmentation accuracy based on manually delineated PVS.

The first benchmark was PINGU. PINGU was trained on six cohorts, including both healthy and clinical populations spanning a broad age range, and was therefore selected as a robust CNN-based state-of-the-art method. For PINGU, segmentation was performed using the PINGU-All_v01 weights, as these were the publicly available weights for download. For PINGU, post-hoc masking of the PVS segmentation with a WMH binary segmentation mask was not suggested or recommended by its developers. Nevertheless, for completeness, all analyses were performed with and without this masking. As it did not appreciably affect any of the findings, results are reported without WMH masking for PINGU.

A second method based on RORPO filtering (RORPO_FSss_) was used as an additional benchmark. The RORPO filter was applied as described above for the semi-automated ground truth dataset, and false positives were minimized using the FreeSurfer SynthSeg tissue mask (RORPO_FSss_), by labelling any voxel within PVS-prone regions (hippocampus, thalamus, white matter and basal ganglia) as PVS, while also excluding any voxel in the WMH binary segmentation mask.

#### 4. Evaluation of segmentation-derived measures of PVS burden using age as a biological benchmark (test6, ADNI, CAHHM datasets)

Segmentation-derived measures of PVS burden were evaluated against age, a well-established biological correlate of PVS burden, to assess the sensitivity of segcsvd and the benchmark tools to an expected source of biological variation. Correlation analyses were conducted between age and two measures of PVS burden (volume and object counts), normalized to total intracranial volume to adjust for head size differences. Reliability was assessed using bias-corrected and accelerated (BCa) bootstrap confidence intervals based on 10,000 resamples. Associations were considered statistically reliable if the confidence interval excluded zero.

#### 5. Robustness of the voxel level segmentation to variation in image contrast and noise (test dataset)

Stability of the voxel level segmentation was evaluated for segcsvd, RORPO, and PINGU under increasing levels of simulated Rician noise and contrast variation, following the methodology introduced in our previous work (Boone et al., 2023).

Noise variation was introduced by applying four levels of Rician noise to the T1-weighted images, with a mean of 1.0 and standard deviations set to 1%, 2%, 3%, and 4% of the maximum voxel intensity (noise factors of 0.01, 0.02, 0.03, and 0.04). Contrast variation was introduced using gamma adjustments, with enhancement factors of 1.1, 1.3, 1.5, and 1.7, and attenuation factors of 0.9, 0.7, 0.5, and 0.3. Each augmentation therefore consisted of five levels, ranging from 0 (no augmentation) to 4 (maximum augmentation), creating a large and diverse dataset to assess robustness to simulated variation in image quality (*n* = 31 images × 3 augmentations × 5 levels = 465).

The noise and contrast augmented T1-weighted images were then segmented using segcsvd, the RORPO filter, and PINGU. For segcsvd, the FreeSurfer Synthseg is a required input, and therefore this was generated after augmentation, providing a true test of performance for segcsvd under these conditions. The FLAIR images used to generate the binary WMH segmentation mask were not subject to this augmentation, as this has been performed previously (Gibson et al., 2024), and the influence of the WMH mask was independently evaluated through ablation-based validation in the current study.

The stability of the voxel level segmentation was assessed across increasing levels of augmentation to quantify agreement relative to each method’s own non-augmented baseline segmentation. This approach enabled a controlled assessment of how variation in image contrast and noise affected segmentation output, independent of baseline accuracy.

#### 6. Robustness of the age-related associations to variation in image contrast and noise (test dataset)

The stability of the associations between age and segmentation-derived measures of PVS burden (volume and object counts) was evaluated for segcsvd, RORPO, and PINGU across increasing levels of augmentation in the test dataset, providing a biologically grounded benchmark of robustness to variation in image contrast and noise that reflects the application of segmentation-derived measures of PVS common in clinical or research analyses.

#### 7. Qualitative review of voxel level segmentation (test, test_6_, ADNI and CAHHM datasets)

In accordance with current guidelines (Pham et al. 2022), a qualitative review was conducted to visually assess the accuracy of segcsvd. Each slice in the test_6_ dataset was examined in a 3D viewer (ITKSnap) for segcsvd, the manual segmentations and the benchmark tools. Manual adjustments to the window level were made to visualize the T1-weighted image under different contrast settings, and agreement between segmented PVS structures and the underlying image features was carefully evaluated. This qualitative approach complemented the quantitative analyses, providing additional insights into segmentation performance differences between segcsvd, the tracers and the benchmark tools.

### Performance metrics

The evaluation framework for segcsvd relied on multiple standard performance metrics calculated at the volume, object, and voxel levels. At the volume level, three metrics were calculated: the intraclass correlation coefficient (ICC), quantifying consistency between segmentation volumes; the Pearson correlation, measuring the strength of the linear association between segmentation volumes without accounting for systematic bias; and the average volume difference (AVD), quantifying absolute relative differences in segmentation volumes.

At the voxel level, four metrics were calculated: the Dice similarity coefficient (DSC), quantifying overall voxel-wise agreement; sensitivity (SNS) quantifying the proportion of true PVS voxels correctly segmented; precision (PRC), quantifying the proportion of segmented voxels that were true PVS; and the normalized surface Dice (NSD), quantifying the spatial alignment of structure boundaries within a 1 mm tolerance and providing an additional boundary-sensitive measure of agreement.

At the object level, three metrics were calculated (DSC, SNS, PRC), with overlap defined at the level of individual PVS objects rather than voxels. These metrics evaluated agreement in detecting the same objects, independent of differences in the extent or volume of those objects.

All metrics were calculated according to established definitions (Maier-Hein et al., 2024).

### Statistical analyses

To assess the reliability of the performance metrics and of correlations between age and segmentation-derived measures of PVS burden (volume and object count), bias-corrected and accelerated (BCa) bootstrap confidence intervals were computed using 10,000 resamples. For the correlations, confidence intervals that excluded zero were considered statistically reliable. Analyses focused on the strength, direction, and consistency of trends across methods and datasets, rather than formal significance testing, reflecting the comparative nature of segmentation performance evaluation.

### Summary of Abbreviations

Abbreviations used for training and validation of segcsvd are summarized in Table 3.

**Table 3.** Summary of the abbreviations used in this work.

| Abbreviation | Description | Use |
| --- | --- | --- |
| RORPO | RORPO-segmented tubular structures | Basis for RORPO<sub>FS</sub> |
| RORPO<sub>FS</sub> | RORPO + FPM with FreeSurfer (FS) mask | Basis for RORPO<sub>GT</sub> |
| RORPO<sub>GT</sub> | RORPO + FPM with FS mask + manual edits | Semi-automated ground truth for model training and ablation validation |
| RORPO<sub>SEED</sub> | RORPO-based centroids + sCSF from FS Synthseg (FSss) | Reference image for manual tracing |
| tracer<sub>1</sub>, tracer<sub>2</sub> | Manual PVS annotations by two human tracers | Manual ground truth segmentations |
| consensus<sub>GT</sub> | Union of tracer<sub>1</sub> and tracer<sub>2</sub> segmentations | Manual ground truth for accuracy validation |
| segcsvd | Current method | Fully automated PVS segmentation |
| segcsvd<sub>edit</sub> | segcsvd after manual removing all false positive voxels | Follow-up analyses of precision in the test<sub>6</sub> dataset |
| RORPO<sub>FSss</sub> | RORPO-segmented “PVS” + FPM with FSss mask | Fully automated RORPO-based benchmark |
| PINGU | PINGU-segmented PVS | Fully automated nnUNet-based benchmark (PINGU-All_v01 weights) |

## Results

### 1. Quantitative assessment of manual editing and tracing efforts

#### Semi-automated ground truth dataset

To assess the extent of manual editing performed to construct the semi-automated ground truth dataset (*n* = 207; RORPO_GT_), agreement between the initial RORPO_FS_ and final edited RORPO_GT_ segmentations was evaluated. Both volume-level agreement (*r* = 0.12 (95% CI: [−0.03, 0.29]) and consistency (ICC = 0.04 [0.00, 0.18]) were low, indicating that substantial manual modifications were made to the initial segmentation volumes.

Voxel-level sensitivity and precision were also calculated, with RORPO_FS_ serving as the reference. In this context, reduced precision reflects the manual addition of true PVS voxels that were absent in RORPO_FS_ (i.e., correction of false negatives), while reduced sensitivity reflects the manual removal of voxels included in RORPO_FS_ (i.e., correction of false positives). Voxel-level sensitivity (SNS = 0.73, [0.70, 0.77]) and precision (PRC = 0.63 [0.59, 0.66]) were both moderate, indicating that a substantial number of both false negative and false positive voxels were present on RORPO_FS_, which were manually corrected to generate RORPO_GT_.

#### Manual ground truth segmentations

To assess the extent of manual tracing performed to generate the manual ground truth segmentations (*n* = 31 × 6 slices), agreement between the unedited RORPO_SEED_ image and the manual segmentations for tracer_1_ and tracer_2_ was assessed.

Volume level correlations were high (tracer_1_: *r* = 0.95, [0.87, 0.98]; tracer_2_: r = 0.93, [0.83, 0.98]), but consistency was poor (tracer_1_: ICC = 0.30, [0.21, 0.40]; tracer2: ICC = 0.29, [0.19, 0.40]), indicating that although larger seed volumes were strongly associated with greater PVS volumes, absolute volume estimates differed substantially from the initial RORPO_SEED_ image.

Voxel level sensitivity and precision were also calculated, with RORPO_SEED_ serving as the reference. Sensitivity was high (tracer_1_: SNS = 0.96, [0.94, 0.98]; tracer_2_: SNS = 0.90, [0.87, 0.93]), indicating that most voxels in RORPO_SEED_ were retained by the tracers. In contrast, precision was low (tracer_1_: PRC = 0.40, [0.38, 0.42]; tracer_2_: PRC = 0.37, [0.35, 0.40]), indicating that a substantial number of additional voxels were traced that were not present on RORPO_SEED_ image.

Together, these results indicate that the final manual segmentations were not strictly determined by RORPO_SEED_ image, reflecting instead substantial independent manual delineation, characterized primarily by the tracing of additional voxels absent from the RORPO_SEED_ image.

#### Consensus manual ground truth segmentations

To evaluate the consistency between the two manually traced segmentations, agreement between tracer_1_ and tracer_2_ was assessed. Volume level correlation was very high (r = 0.98, [0.96, 1.00]), with excellent consistency (ICC = 0.98, 95% CI: [0.96, 1.00]), and strong voxel level agreement (DSC = 0.84, [0.82, 0.86]). Given the high degree of agreement in the manual annotations between the two tracers, a consensus manual ground truth segmentation (consensus_GT_) was created that included the union of all voxels identified by either tracer, prioritizing sensitivity to all manually segmented PVS voxels.

### 2. Validation of segcsvd model design

The performance of segcsvd was first validated using the test dataset (*n* = 31) and measured against the semi-automated ground truth segmentations (RORPO_GT_), which served as the training target for segcsvd. The full model was compared to ablated variants with key components removed: WMH masking (segcsvd_no_wmh_), test-time intensity augmentation (segcsvd_no_aug_), epoch ensembling (segcsvd_1epoch_), and Tversky loss ensembling (segcsvd_1loss_) (Table 4).

**Table 4.** Performance metrics for segcsvd and ablated variants on the test dataset, evaluated with respect to the semi-automated reference (RORPO_GT_). Ablated variants exclude key components of the full model: WMH segmentation mask (segcsvd_no_wmh_), test-time intensity augmentation (segcsvd_no_aug_), epoch ensembling (segcsvd_1epoch_), and Tversky loss ensembling (segcsvd_1loss_). Values in square brackets denote the 95% bootstrap estimated confidence intervals.

| Performance Metrics |  |  |  |  |  |  |  |  |  |
| --- | --- | --- | --- | --- | --- | --- | --- | --- | --- |
| BG PVS | Volume | Voxel |  |  |  | Volume | Object |  |  |
|  | Corr | Dice | Sensitivity | Precision | NSD | AVD | Dice | Sensitivity | Precision |
| **segcsvd** | 0.88<br>[0.65, 0.93] | 0.67<br>[0.63, 0.70] | 0.81<br>[0.75, 0.84] | 0.60<br>[0.54, 0.63] | 0.84<br>[0.81, 0.86] | 0.53<br>[0.37, 0.87] | 0.71<br>[0.68, 0.74] | 0.79<br>[0.73, 0.83] | 0.68<br>[0.63, 0.72] |
| **segcsvd<sub>no_wmh</sub>** | 0.89<br>[0.76, 0.93] | 0.67<br>[0.63, 0.70] | 0.80<br>[0.74, 0.84] | 0.61<br>[0.55, 0.64] | 0.84<br>[0.81, 0.86] | 0.49<br>[0.34, 0.80] | 0.72<br>[0.69, 0.74] | 0.78<br>[0.73, 0.83] | 0.70<br>[0.66, 0.73] |
| **segcsvd<sub>no_aug</sub>** | 0.88<br>[0.68, 0.93] | 0.68<br>[0.63, 0.71] | 0.79<br>[0.73, 0.83] | 0.62<br>[0.57, 0.66] | 0.84<br>[0.82, 0.86] | 0.47<br>[0.33, 0.80] | 0.72<br>[0.69, 0.75] | 0.78<br>[0.72, 0.83] | 0.70<br>[0.66, 0.74] |
| **segcsvd<sub>1epoch</sub>** | 0.88<br>[0.65, 0.93] | 0.68<br>[0.63, 0.71] | 0.76<br>[0.71, 0.81] | 0.64<br>[0.59, 0.68] | 0.84<br>[0.81, 0.86] | 0.40<br>[0.28, 0.65] | 0.72<br>[0.68, 0.74] | 0.75<br>[0.69, 0.79] | 0.73<br>[0.68, 0.77] |
| **segcsvd<sub>1loss</sub>** | 0.85<br>[0.51, 0.92] | 0.67<br>[0.63, 0.70] | 0.76<br>[0.71, 0.80] | 0.62<br>[0.56, 0.66] | 0.83<br>[0.80, 0.85] | 0.42<br>[0.28, 0.73] | 0.71<br>[0.67, 0.73] | 0.76<br>[0.70, 0.80] | 0.69<br>[0.64, 0.73] |
| NBG PVS | Volume | Voxel |  |  |  | Volume | Object |  |  |
|  | Corr | Dice | Sensitivity | Precision | NSD | AVD | Dice | Sensitivity | Precision |
| **segcsvd** | 0.99<br>[0.97, 1.00] | 0.65<br>[0.62, 0.68] | 0.78<br>[0.74, 0.81] | 0.57<br>[0.53, 0.60] | 0.80<br>[0.78, 0.83] | 0.46<br>[0.35, 0.61] | 0.67<br>[0.64, 0.69] | 0.76<br>[0.71, 0.80] | 0.62<br>[0.57, 0.66] |
| **segcsvd<sub>no_wmh</sub>** | 0.97<br>[0.86, 1.00] | 0.56<br>[0.51, 0.61] | 0.79<br>[0.75, 0.82] | 0.46<br>[0.40, 0.52] | 0.74<br>[0.71, 0.78] | 1.02<br>[0.70, 1.38] | 0.63<br>[0.59, 0.66] | 0.77<br>[0.72, 0.80] | 0.56<br>[0.50, 0.61] |
| **segcsvd<sub>no_aug</sub>** | 0.99<br>[0.97, 1.00] | 0.66<br>[0.63, 0.69] | 0.75<br>[0.70, 0.78] | 0.60<br>[0.56, 0.63] | 0.81<br>[0.78, 0.83] | 0.35<br>[0.26, 0.47] | 0.67<br>[0.64, 0.70] | 0.72<br>[0.67, 0.77] | 0.65<br>[0.61, 0.69] |
| **segcsvd<sub>1epoch</sub>** | 0.99<br>[0.97, 1.00] | 0.65<br>[0.63, 0.68] | 0.75<br>[0.71, 0.79] | 0.59<br>[0.56, 0.63] | 0.80<br>[0.78, 0.83] | 0.36<br>[0.28, 0.48] | 0.67<br>[0.64, 0.70] | 0.72<br>[0.68, 0.77] | 0.65<br>[0.60, 0.69] |
| **segcsvd<sub>1loss</sub>** | 0.99<br>[0.96, 1.00] | 0.64<br>[0.61, 0.67] | 0.74<br>[0.70, 0.77] | 0.58<br>[0.54, 0.62] | 0.79<br>[0.77, 0.82] | 0.37<br>[0.26, 0.49] | 0.66<br>[0.62, 0.69] | 0.72<br>[0.68, 0.77] | 0.63<br>[0.58, 0.67] |

For BG PVS, segcsvd showed moderately strong voxel-level agreement with RORPO_GT_ (DSC = 0.67, 95% CI: [0.63, 0.70]), characterized by high sensitivity (SNS = 0.81, [0.75, 0.84]) and lower precision (PRC = 0.60, [0.54, 0.63]). Similar results were observed both at the object-level and for NBG PVS, with differences in these scores not exceeding 4% of those observed for BG PVS. Together, these results reflect a consistent trade-off for segcsvd favoring sensitivity over precision relative to RORPO_GT_.

Omission of the WMH segmentation mask (segcsvd_no_wmh_) had a minimal impact on performance for BG PVS, with voxel and object-level metrics differing by approximately 1% relative to segcsvd. This omission had a larger impact for NBG PVS, where voxel-level precision decreased by 10% and object-level precision decreased by 6%, indicating a notable increase in false positives when the WMH mask was excluded.

Omission of test-time intensity augmentation (segcsvd_no_aug_), and reduced ensembling (segcsvd_1epoch_, segcsvd_1loss_) had a minimal impact on performance, resulting in a modest shift in the sensitivity-precision trade-off at both the voxel and object level for BG and NBG PVS, with sensitivity decreasing and precision increasing by approximately 1-5% relative to segcsvd.

### 3. Validation of segmentation accuracy for segcsvd and two benchmark tools

The performance of segcsvd and the two benchmark tools (PINGU, RORPO_FSss_) was evaluated using the test_6_ dataset by measuring agreement with the manual ground truth segmentations (consensus_GT_) (Table 5, Figure 3).

**Table 5.** Performance metrics for segcsvd and two alternative methods (PINGU, RORPO_FSss_) on the test_6_ dataset, evaluated with respect to the consensus_GT_ manual ground truth reference. Values in square brackets denote the 95% bootstrap estimated confidence intervals.

| Performance Metrics |  |  |  |  |  |  |  |  |  |
| --- | --- | --- | --- | --- | --- | --- | --- | --- | --- |
| BG PVS | Volume | Voxel |  |  |  |  | Object |  |  |
|  | Corr | Dice | Sensitivity | Precision | NSD | AVD | Dice | Sensitivity | Precision |
| segcsvd | 0.87<br>[0.70, 0.92] | 0.65<br>[0.61, 0.70] | 0.80<br>[0.75, 0.85] | 0.57<br>[0.52, 0.63] | 0.83<br>[0.73, 0.87] | 0.55<br>[0.40, 0.80] | 0.78<br>[0.73, 0.82] | 0.80<br>[0.74, 0.85] | 0.78<br>[0.73, 0.83] |
| PINGU | 0.51<br>[0.01, 0.77] | 0.60<br>[0.52, 0.65] | 0.59<br>[0.51, 0.64] | 0.70<br>[0.60, 0.77] | 0.75<br>[0.66, 0.81] | 0.34<br>[0.27, 0.43] | 0.64<br>[0.55, 0.69] | 0.60<br>[0.53, 0.68] | 0.79<br>[0.68, 0.86] |
| RORPO<sub>FSSs</sub> | 0.92<br>[0.85, 0.96] | 0.74<br>[0.71, 0.78] | 0.81<br>[0.78, 0.84] | 0.70<br>[0.65, 0.75] | 0.88<br>[0.75, 0.92] | 0.27<br>[0.19, 0.36] | 0.91<br>[0.89, 0.93] | 0.88<br>[0.83, 0.91] | 0.95<br>[0.93, 0.97] |
| NBG PVS | Volume | Voxel |  |  |  |  | Object-Level |  |  |
|  | Corr | Dice | Sensitivity | Precision | NSD | AVD | Dice | Sensitivity | Precision |
| segcsvd | 0.95<br>[0.88, 0.98] | 0.60<br>[0.54, 0.64] | 0.67<br>[0.60, 0.73] | 0.57<br>[0.52, 0.62] | 0.77<br>[0.71, 0.81] | 0.39<br>[0.30, 0.49] | 0.67<br>[0.63, 0.71] | 0.67<br>[0.61, 0.72] | 0.71<br>[0.66, 0.76] |
| PINGU | 0.85<br>[0.70, 0.93] | 0.52<br>[0.45, 0.57] | 0.63<br>[0.56, 0.70] | 0.49<br>[0.42, 0.55] | 0.69<br>[0.61, 0.73] | 0.71<br>[0.48, 1.18] | 0.56<br>[0.49, 0.61] | 0.61<br>[0.54, 0.67] | 0.57<br>[0.50, 0.63] |
| RORPO<sub>FSSs</sub> | 0.95<br>[0.88, 0.98] | 0.59<br>[0.50, 0.64] | 0.61<br>[0.53, 0.67] | 0.59<br>[0.51, 0.66] | 0.73<br>[0.64, 0.78] | 0.42<br>[0.22, 0.83] | 0.66<br>[0.56, 0.72] | 0.69<br>[0.60, 0.75] | 0.67<br>[0.57, 0.75] |

**Figure 3.**
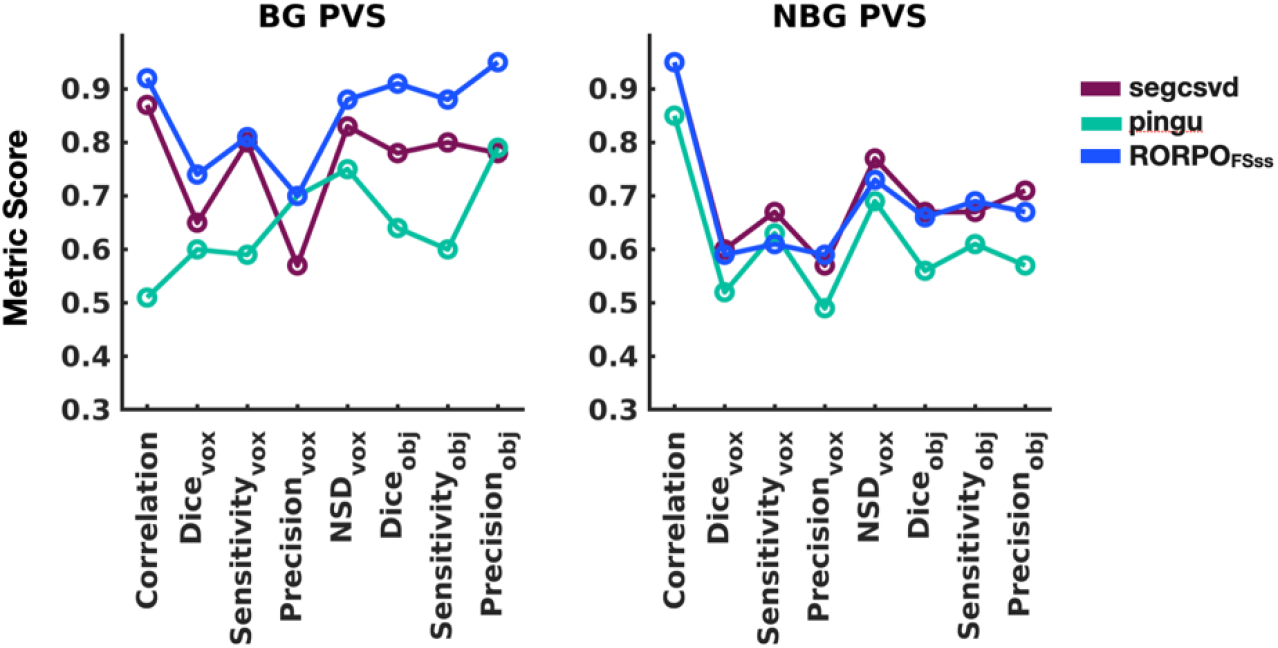
Performance metrics for segcsvd and two benchmark methods (PINGU, RORPO_FSss_) for the test_6_ dataset, evaluated with respect to the consensus_GT_ manual ground truth reference. Means are plotted to illustrate overall trends across methods. In general, all methods performed better for BG PVS than NBG PVS. Overall, PINGU exhibited the lowest performance for both BG and NBG PVS across nearly all metrics. RORPOFS_ss_ generally showed the strongest performance for BG PVS, while segcsvd performed marginally better for NBG PVS across most metrics. For segcsvd, precision for BG PVS was low, but was accompanied by particularly high sensitivity, reflecting a trade-off that favored more inclusive segmentation at the expense of false positives

For BG PVS, segcsvd showed strong performance at the object level (DSC = 0.78 [0.73, 0.82]; SNS = 0.80 [0.74, 0.85]; PRC = 0.78 [0.73, 0.83]), higher than PINGU (DSC = 0.64 [0.55, 0.69], SNS = 0.60 [0.53, 0.68], PRC = 0.79 [0.68, 0.86]), but lower than RORPO_FSss_ (DSC = 0.91 [0.89, 0.93], SNS = 0.88 [0.83, 0.91], PRC = 0.95 [0.93, 0.97]). At the voxel level, sensitivity remained high for segcsvd (SNS = 0.80 [0.75, 0.85]), comparable with RORPO_FSss_ (SNS = 0.81 [0.78, 0.84], and higher than PINGU (SNS = 0.59 [0.51, 0.64]). In contrast, voxel level precision for segcsvd (PRC = 0.57 [0.52, 0.63]) was reduced compared to both PINGU (PRC = 0.70 [0.60, 0.77]) and RORPO_FSss_ (PRC = 0.70 [0.65, 0.75]). Together, high object- and voxel-level sensitivity, combined with high object-level precision and reduced voxel-level precision, indicates that segcsvd accurately detects BG PVS, but tends to segment them with more liberal boundaries, resulting in larger delineations relative to the manual ground truth.

For non-BG PVS, all methods showed reduced performance compared to BG PVS across most metrics, indicating increased difficulty of segmenting PVS in this region (Figure 3). Object-level performance was higher for segcsvd (DSC = 0.67 [0.63, 0.71], SNS = 0.67 [0.61, 0.72], PRC = 0.57 [0.50, 0.63]) and RORPO_FSss_ (DSC = 0.66 [0.56, 0.72], SNS = 0.69 [0.60, 0.75, PRC = 0.67 [0.57, 0.75]) compared to PINGU (DSC = 0.66 [0.56, 0.72], SNS = 0.61 [0.54, 0.67], PRC = 0.57 [0.50, 0.63]. Voxel-level performance was higher for segcsvd (DSC = 0.60 [0.54, 0.64], SNS = 0.67 [0.60, 0.73], PRC = 0.57 [0.52, 0.62], NSD = 0.77 [0.71, 0.81]), particularly in terms of sensitivity and surface distance, compared to PINGU (DSC = 0.52 [0.45, 0.57], SNS = 0.63 [0.56, 0.70], PRC = 0.49 [0.42, 0.55], NSD = 0.69 [0.61, 0.73]) and RORPO_FSss_ (DSC = 0.59 [0.50, 0.64], SNS = 0.61 [0.53, 0.67], PRC = 0.59 [0.51, 0.66], NSD = 0.73 [0.64,0.78]), highlighting a consistent performance advantage for segvcsvd in this more challenging region.

#### segcsvd_edit_

Given that segcsvd exhibited generally strong performance despite lower voxel-level precision for BG PVS, a follow-up analysis was conducted to assess whether precision could be improved through manual correction. The segcsvd segmentation was manually edited to remove all false positive non-PVS voxels, and precision was again calculated relative to the manual ground truth segmentation (concensus_GT_).

This manual editing resulted in only a modest improvement in precision of approximately 3% for segcsvd_edit_ compared to segcsvd. For BG PVS, voxel-level precision increased from 0.57 [0.52, 0.63] to 0.60 [0.54, 0.66], and object-level precision increased from 0.78 [0.73, 0.83] to 0.81 [0.76, 0.86]. For non-BG PVS, voxel-level precision increased from 0.57 [0.52, 0.62] to 0.60 [0.53, 0.65], while object-level precision increased from 0.71 [0.66, 0.75] to 0.73 [0.65, 0.79]. Thus, voxel-level precision remained low for segcsvd even after removing all false-positive PVS voxels, indicating that the low voxel-level precision was not driven by true false-positive segmentation errors that could be manually corrected.

### 4. Evaluation of segmentation-derived measures of PVS burden using age as a biological benchmark

The strength of the associations between age and segmentation-derived measures of PVS burden (volumes and object counts) was examined for segcsvd, RORPO_FSss_ and PINGU in the test_6_, CAHHM, and ADNI datasets. For the test_6_ dataset with manual ground truth segmentations, concensus_GT_ and segcsvd_edit_ were also included in this analysis.

#### test_6_ dataset

Only segcsvd and segcsvd_edit_ consistently detected statistically reliable positive associations with age for all eight of the examined associations in the test_6_ dataset, as indicated by confidence intervals that did not include zero (Figure 4 A-D). For BG PVS volume (Figure 4A), the strength of the association with age was nearly identical for segcsvd (*r* = 0.41 [0.04, 0.60]) and segcsvd_edit_ (*r* = 0.40 [0.02, 0.59]), indicating that manual editing to remove false positives had minimal impact on the observed association with age. Slightly weaker correlations were observed for concensus_GT_ (*r* = 0.34 [−0.06, 0.59]) and RORPO_FSss_ (*r* = 0.33 [−0.06, 0.58]). In contrast, a negative association with age was observed for PINGU (*r* = −0.28 [−0.57, 0.05]).

**Figure 4.**
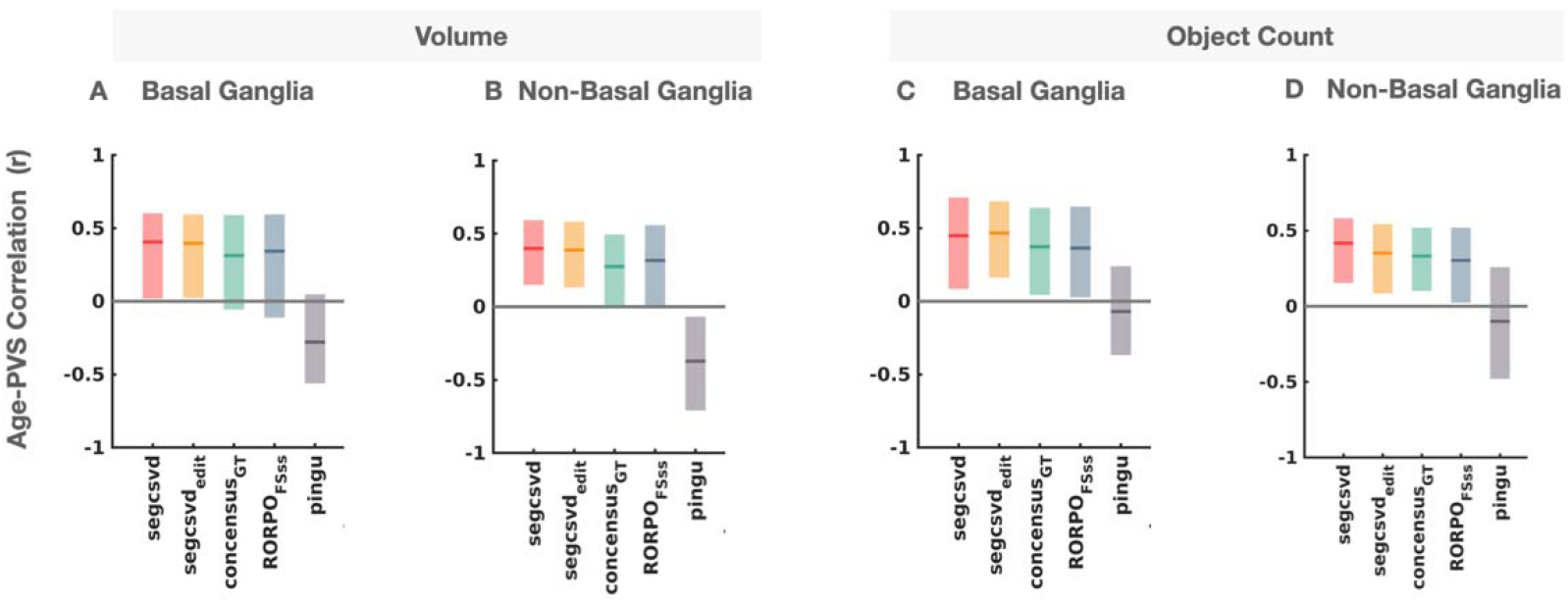
Associations between age and segmentation-derived measures of PVS burden (volumes and object counts) for the test_6_ dataset. Shaded areas indicate the Pearson correlation coefficient with 95% bootstrap estimated confidence intervals. Only segcsvd and segcsvd_ed t_ detected statistically reliable positive associations with age in all cases (A-D), as indicated by confidence intervals that did not include zero.

A similar pattern of effects was observed for all other PVS measures (Figure 4 B-D), with PINGU showing both the greatest variability in the strength of the correlation across measures and unexpected negative correlations with age, particularly for PVS volumes. The concensus_GT_ showed greater reliability at the object level than at the voxel level, as indicated by confidence intervals that did not include zero.

#### CAHHM and ADNI datasets

Overall, segcsvd was more effective than both RORPO_FSss_ and PINGU at detecting age-related associations with both PVS volume and counts in both the CAHHM and ADNI datasets. Specifically, segcsvd often identified stronger positive associations with age and was the only method to consistently detect statistically reliable associations in all eight cases, as indicated by confidence intervals that did not include zero (Figure 5).

**Figure 5.**
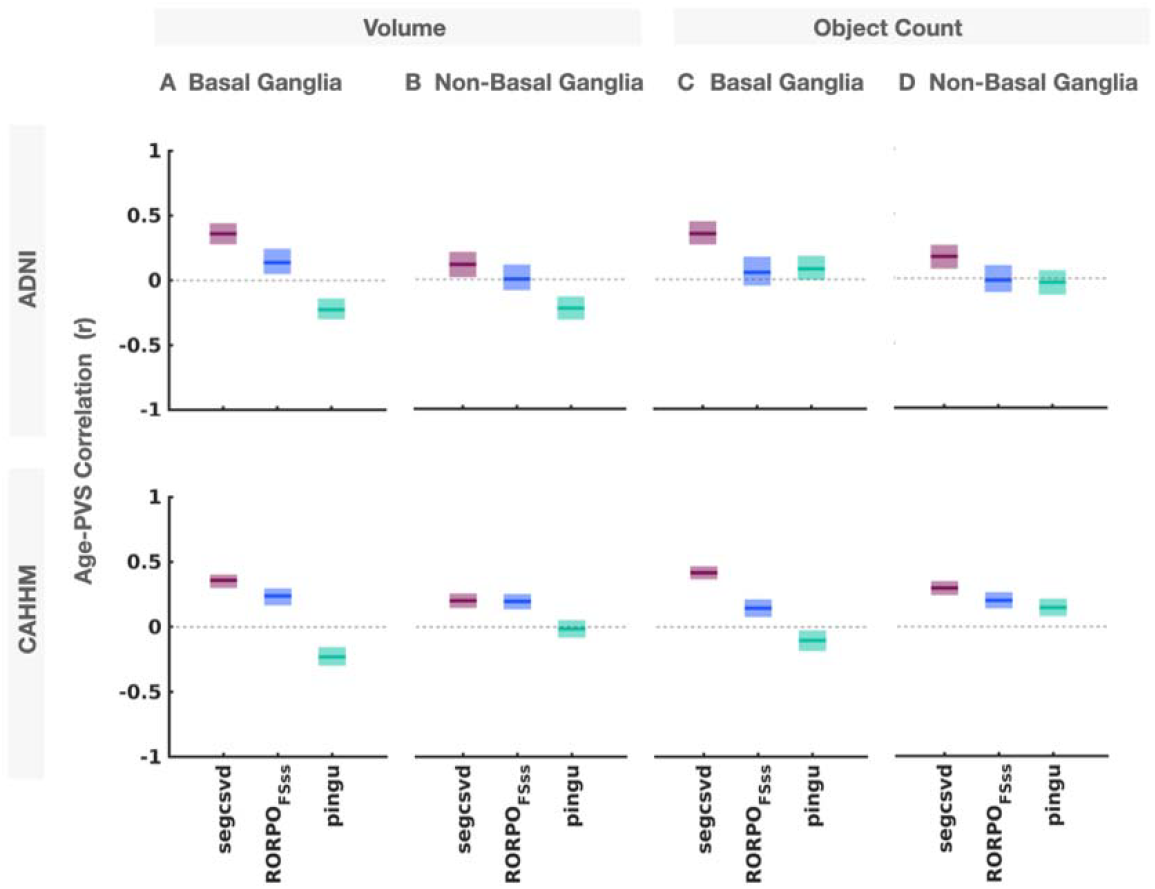
Associations between age and segmentation-derived measures of PVS burden (volumes and object counts) for the ADNI and CAHHM cohorts. Shaded areas indicate the Pearson correlation coefficient with 95% bootstrap estimated confidence intervals. Compared to both RORPO_FSss_ and PINGU, segcsvd tended to identify stronger positive associations with age and was the only method to detect statistically reliable associations for all eight of the examined associations, as indicated by confidence intervals that did not include zero.

For basal ganglia PVS volume, all three methods detected reliable associations with age. These associations were in the expected positive direction for both segcsvd (ADNI: *r* = 0.36; [0.28 0.44]; CAHHM: *r* =0.36 [0.31, 0.41]) and RORPO_FSss_ (ADNI: *r* = 0.14 [0.05, 0.24]; CAHHM: *r* = 0.24 [0.17, 0.30]). In both the ADNI and CAHHM cohorts, the confidence intervals for segcsvd and RORPO_FSss_ did not overlap, indicating reliably stronger associations for segcsvd. In contrast, this association was statistically reliable but in the unexpected negative direction for PINGU (ADNI: *r* = −0.22 [−0.30, −0.14]; CAHHM: *r* = −0.23 [−0.16, −0.30]).

Across all eight examined associations, several broad trends emerged. Associations tended to be more variable for PINGU, which showed both statistically reliable positive (*n* = 1) and negative associations (*n* = 5); whereas segcsvd tended to show stronger and more reliable positive associations than RORPO_FSss_, particularly for BG PVS volumes and object counts.

### 5. Robustness of the voxel-level segmentation to variation in image contrast and noise (out-of-distribution data)

The stability of the segmentation output was assessed for segcsvd, RORPO_FSss_, and PINGU using the test dataset under increasing levels of contrast enhancement, contrast attenuation, and Rician noise, with performance metrics calculated relative to each method’s baseline output (i.e., without augmentation); evaluating the stability of performance independent of baseline accuracy (Figure 6).

**Figure 6.**
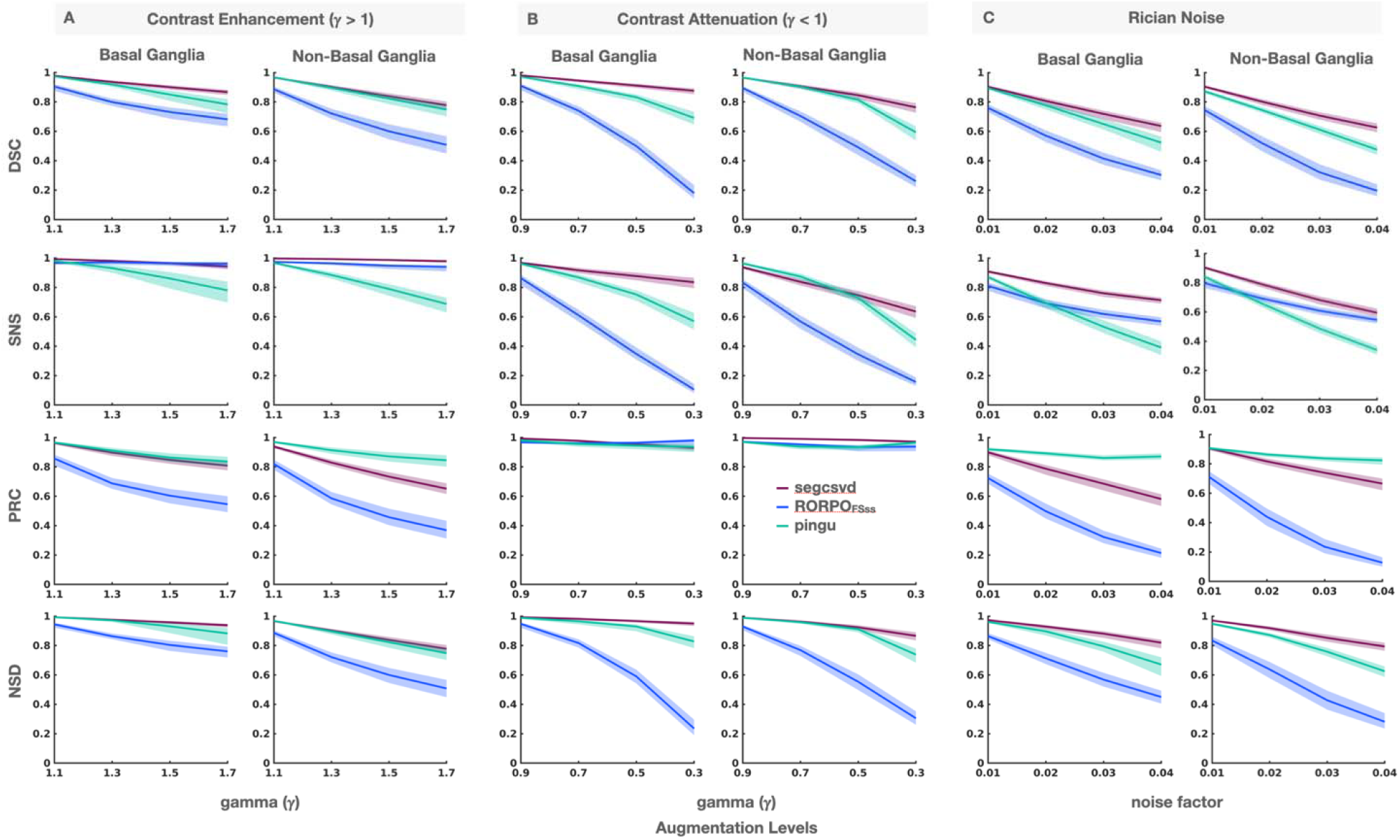
Performance metrics under increasing levels of contrast and noise augmentation for the test dataset. Metrics were calculated relative to the original, non-augmented data, assessing segmentation stability independent of accuracy (i.e. agreement with semi-automated or manual ground truth references). Shaded areas indicate mean values and 95% bootstrap-estimate confidence intervals. Overall, the CNN-based methods (segcsvd, PINGU) exhibited greater stability in the performance metrics, reflecting greater robustness to simulated variation in image contrast and noise.

Overall, the CNN-based methods (segcsvd, PINGU) demonstrated greater robustness to increasing levels of contrast and noise augmentation compared to the RORPO-based method (RORPO_FSss_). Dice scores for RORPO_FSss_ declined sharply, particularly under contrast attenuation and Rician noise, where they ranged between approximately 0.2 and 0.3 at the highest augmentation levels. In contrast, PINGU and segcsvd showed more gradual declines, maintaining Dice scores above approximately 0.6 (segcsvd) or 0.5 (PINGU) across all augmentation levels, indicating greater overall stability in their voxel-level segmentation output.

While the Dice scores for segcsvd and PINGU were generally comparable across all augmentation levels for BG and NBG PVS, differences in sensitivity and precision were observed. For PINGU, sensitivity declined consistently with increasing levels of augmentation, ranging between approximately 0.4 and 0.8 at the highest augmentation levels. In contrast, segcsvd maintained relatively higher sensitivity, ranging between approximately 0.7 and 1.0 at the highest levels. For segcsvd, this was generally accompanied by a greater decline in precision, which ranged between approximately 0.6 and 1.0 across the highest levels, compared to PINGU, which ranged between approximately 0.8 and 1.0.

### 6. Robustness of the age-related associations to variation in image contrast and noise

The stability of age-related associations with PVS measures (volumes and object counts) was evaluated using the test dataset for segcsvd, RORPO_FSss_ and PINGU across increasing levels of contrast and noise augmentation (Figure 7).

**Figure 7.**
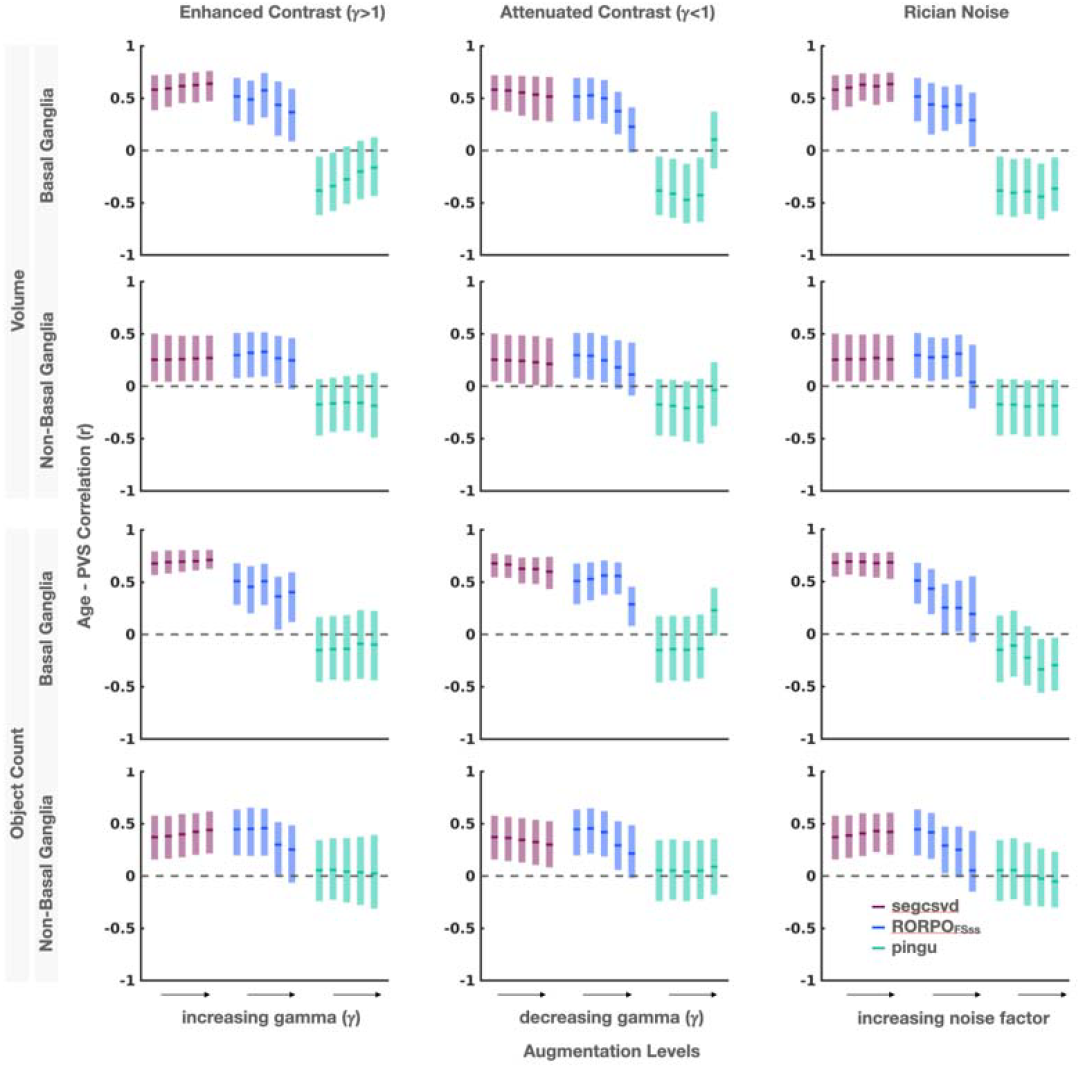
Strength of the age-related associations under increasing levels of contrast and noise augmentation in the test dataset. Shaded areas indicated the Pearson correlation coefficient and 95% bootstrap estimated confidence intervals. Correations are shown for five augmentation levels (0: no augmentation, to 4: maximum augmentation) for each method. Overall, segcsvd exhibited greater stability in these correlations across increasing levels of augmentation compared to both RORPO_FSss_ and PINGU.

For segcsvd, age-related correlations were highly stable, with only minimal changes in their strength across five increasing levels of contrast and noise augmentation (Δ*r* < 0.1). These changes exhibited a consistent trend, where enhancement and noise augmentation resulted in largely linear increases in correlation strength, while increasing contrast attenuation resulted in largely linear decreases in correlation strength.

For RORPO_FSss_, age-related correlations were less stable, with larger changes in strength observed across the five augmentation levels (Δr < 0.4). These changes followed a consistent pattern, with all augmentation types resulting in largely linear decreases in correlation strength.

For PINGU, age-related correlations exhibited the greatest variability across augmentation levels. Confidence intervals were generally wider, and changes in correlation strength lacked a consistent pattern. While some measures showed increasing or decreasing correlations with augmentation, others remained relatively stable, indicating no systematic directionality in the response to changes in image contrast and noise.

### 7. Qualitative evaluation of segcsvd segmentation performance

The segmentation outputs from the tracers, segcsvd, RORPO_FSss_ and PINGU were examined visually for the test_6_ dataset (Figure 8). Compared to the tracers, segcsvd consistently delineated larger PVS structures, particularly in the basal ganglia for individuals with high PVS burden. The larger delineation of PVS objects by segcsvd aligned closely with the T1 image when visualized using a compressed intensity range to enhance the appearance of hypointense voxels (Figure 9). PINGU showed promising but generally inconsistent performance (Figure 8). While PINGU appeared to better capture the shape or extent of PVS in some cases, particularly in white matter, this was accompanied by frequent missed PVS, particularly in the basal ganglia. Occasional false-positive inclusions of non-PVS hypointensities within WMH or other GM structures were also observed. RORPO_FSss_, showed good agreement with the tracers, particularly in the extent or size of PVS, with most errors arising from imperfect masking (Figure 8).

**Figure 8.**
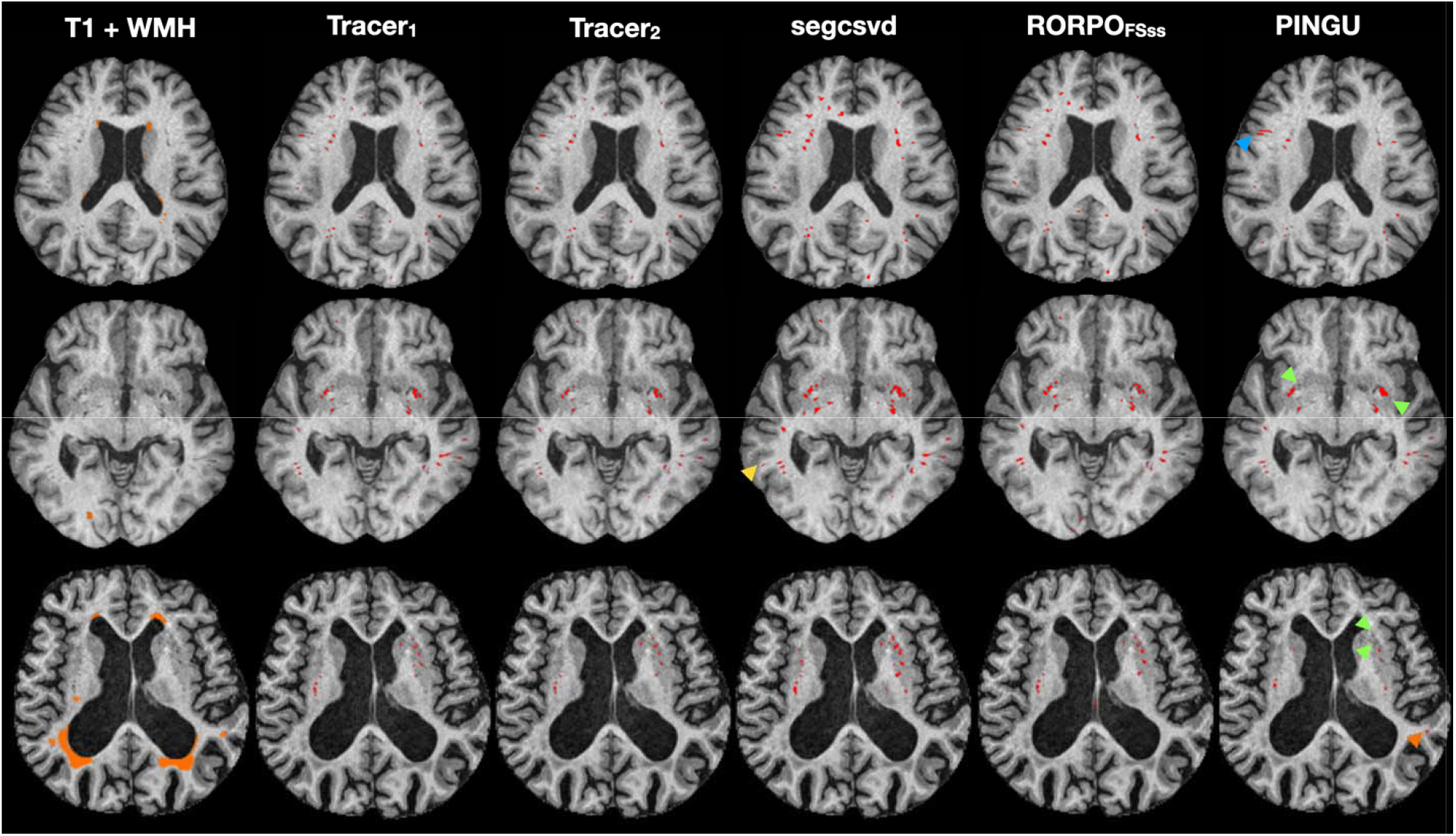
Qualitative comparison of segmentation performance for segcsvd, the tracers and the benchmark tools in three representative slices from the test_6_ dataset. The FLAIR-based WMH segmentation overlaid in orange on the T1-weighted image (column 1). Compared to the tracers, segcsvd occasionally captured true PVS missed by both tracers (e.g., yellow arrow), and generally showed good agreement, with a tendency to segment PVS with greater extent. Errors for RORPO_FSss_ often arose from imperfect false positive minimization. PINGU showed more variable performance, occasionally capturing the extent or shape of PVS well, particularly in white matter (e.g., blue arrow), but frequently missing PVS in the basal ganglia (green arrows), especially in older or more clinically advanced cases (bottom row), while also showing occasional false positives within WMH (e.g., orange arrow) or other GM structures.

**Figure 9.**
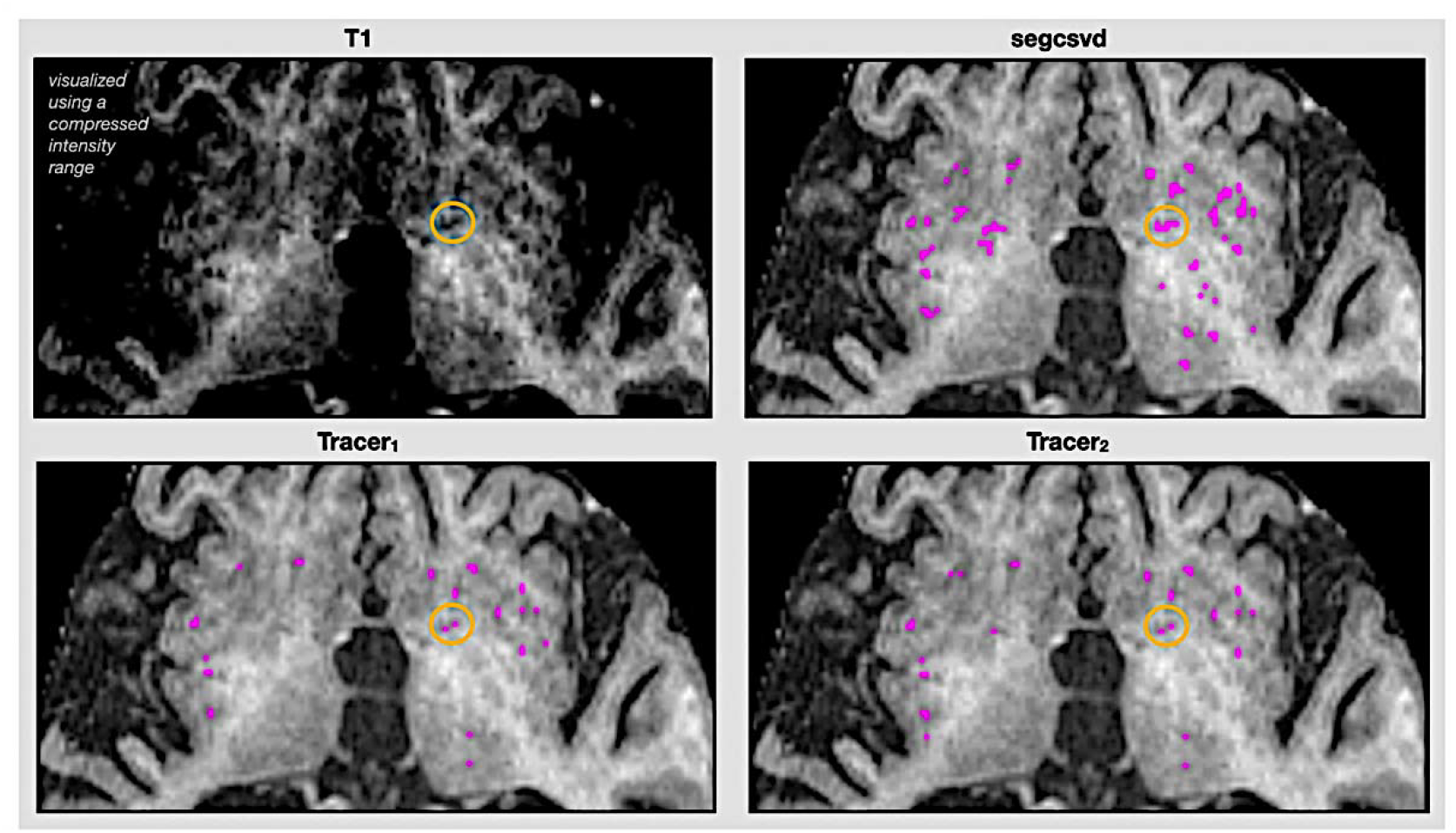
Typical segmentation differences between segcsvd and the tracers in the basal ganglia for an individual with high PVS burden. The larger delineation of PVS extent by segcsvd aligned more closely with T1 image when visualized using a compressed intensity range to enhance the appearance of hypointense voxels (e.g., circled area).

## Discussion

This study presents segcsvd, a fully automated CNN-based PVS segmentation method for T1-weighted images. The model incorporates anatomical priors from other validated segmentation methods and is trained on a large, multisite dataset to promote generalizability. Its performance was compared to existing benchmark tools using a comprehensive evaluation framework encompassing ablation studies, validation against manual ground truth, sensitivity to known associations with age, and robustness to image variation.

### Accuracy relative to manual ground truth

Accuracy was validated against manual ground truth using the test_6_ dataset, where several broad trends emerged. First, performance was consistently lower for non-basal ganglia PVS across all methods, with reduced Dice scores, sensitivity, and precision at both the voxel and object levels compared to basal ganglia PVS. While scores for segcsvd and RORPO_FSss_ were approximately 10% higher than those for PINGU, the overall pattern suggests that non-basal ganglia PVS remains a more challenging target for PVS segmentation on T1-weighted images. For CNN-based methods, training on even larger datasets may be necessary to better learn features that differentiate PVS from anatomically or artifactually confounding structures in this region. For filter-based methods, more sophisticated false-positive minimization strategies may be needed, as false positives may occur more frequently in this region, due to its larger size and greater heterogeneity in structural and intensity profiles.

Second, PINGU generally showed lower performance compared to both segcsvd and RORPO_FSss_ across all performance metrics. The observed performance scores for PINGU were broadly consistent with those reported in the original publication, indicating that the tool performed largely as designed when applied to our test dataset and evaluated against our manual ground truth. One exception was noted, where the volume correlation of basal ganglia PVS was 0.51 in our test data, which was approximately 10% below the lower bound of the reported range (0.77⍰±⍰0.14) across test datasets in the original publication. This discrepancy may reflect differences in the clinical characteristics of the test data used in this study. Although this dataset was randomly partitioned, it included individuals with more advanced neurodegenerative changes, as indicated by a high percentage of ventricular CSF (%VCSF = 3.2⍰±⍰1.6), potentially posing a more challenging test of generalization. These findings underscore the importance of continued development and validation across clinically and demographically diverse populations for CNN-based segmentation tools.

Third, for basal ganglia PVS, object-level performance for segcsvd was high (DSC = 0.78; SNS = 0.80; PRC = 0.78), while voxel-level performance was lower, with sensitivity remaining high (SNS = 0.80) but precision decreasing (PRC = 0.57). The decrease in voxel-level precision was likely driven, in part, by deliberate design choices, including test-time intensity augmentation and model ensembling, intended to enhance the detection of subtle PVS on T1-weighted images. Ablation studies confirmed a consistent trade-off, with increased sensitivity accompanied by decreased precision. However, the magnitude of this effect was relatively modest, suggesting additional factors contributed to the lower voxel-level precision. One such factor may be a more liberal segmentation tendency, where segcsvd may have more closely followed image intensity patterns and detected subtle signal variations not consistently captured in manual annotations. Supporting this interpretation, post hoc manual editing of segcsvd to remove all false positives voxels (segcsvd_edit_) resulted in only a modest increase in voxel-level precision (approximately 3%). The presence of high object-level precision, together with low voxel-level precision that persists even after thorough manual corrections, suggests a systematic difference in segmentation criteria rather than random oversegmentation of object boundaries. Visualization of the T1 image using a compressed intensity range revealed close alignment with segcsvd segmentations, further supporting this interpretation.

### Age as biological benchmark

In the test_6_ dataset, correlations between age and segmentation-derived measures of PVS burden (volume and object counts) were strong, statistically reliable, and in the expected positive direction for segcsvd. Importantly, these associations were largely unchanged (Δr < 0.1) after manually removing all false positive voxels (segcsvd_edit_),indicating that the observed effects were not driven by incorrect inclusion of non-PVS voxels that were themselves associated with age (e.g., lacunes).

The strength of these associations was stronger for segcsvd than for the consensus manual ground truth segmentation, which showed statistically reliable associations with age only at the object level but not at the voxel level. This suggests that the presence of PVS was more consistently detected than precisely delineated in the manual ground truth dataset, potentially reflecting limitations of manual annotation in consistently tracing small structures. In contrast, PINGU exhibited more variable and negative correlations with age, which were inconsistent with both the consensus manual ground truth segmentation and expected biological patterns.

### Robustness of the voxel-level segmentation to variation in contrast and noise

Overall, both segcsvd and PINGU demonstrated greater stability in their voxel level segmentation under simulated variation in image contrast and noise compared RORPO_FSss_. These results highlight a key limitation of filter-based methods, which often require manual parameter tuning to account for differences in image contrast and quality across datasets. In contrast, while CNN-based approaches still face challenges in generalization, these findings underscore their potential as a more robust and scalable solution across diverse imaging conditions.

### Robustness of associations between age and PVS burden to variation in contrast and noise

In general, segcsvd demonstrated remarkable consistency in the strength of age-related associations with PVS burden across increasing levels of simulated variation in contrast and noise variation compared to both benchmark tools. Moreover, a small upward trend in the strength of the correlations was observed for segcsvd with increasing levels of contrast enhancement and noise. These findings may be explained, in part, by the phenomenon of stochastic resonance, where the addition of a small amount of noise can enhance detection sensitivity by elevating sub-threshold signals, such as voxel intensity, above a detection threshold (McDonnel And Abbot, 2009). Likewise, contrast enhancement amplifies the intensity of subtle PVS signals, which may similarly improve the detection of true PVS. These findings provide further support in favor of prioritizing sensitivity over precision in this task, as this approach may enhance the detection of biologically relevant associations for small, subtle structures like PVS.

More broadly, these results underscore the challenges inherent in validating segmentation tools. In addition to potential biases and errors in the reference segmentations used as ground truth, other factors, such as image noise, may reduce voxel level segmentation consistency without diminishing, and in some cases, potentially enhancing, associations with biologically relevant variables such as age. These considerations reinforce the value of a comprehensive evaluation strategy that assesses segmentation accuracy alongside biological benchmarks, rather than relying solely on standard performance metrics.

### Caveats and Limitations

While segcsvd demonstrated strong performance across multiple evaluation criteria, several caveats and limitations should be noted. First, a reference image (RORPO_SEED_) was used to enhance the accuracy of the manual ground truth dataset. Although this introduced potential detection bias toward RORPO-segmented structures, agreement analyses confirmed that substantial independent tracing was performed by each tracer beyond the initial seed voxels. Moreover, given that RORPO is widely used for PVS quantification and has been independently validated using in-silico phantoms (Duarte Coello, 2024), this strategy was designed to improve segmentation accuracy by providing tracers with a minimal visual reference derived from a well-established method—an approach motivated by the known difficulty of accurately delineating PVS through manual tracing alone (Pham et al., 2022).

Second, the manual ground truth dataset was limited in size. While the slices were selected to maximize anatomical diversity, the small sample size reflects the practical constraints of manually annotating subtle and spatially diffuse structures, such as PVS.

Third, while informative, the use of age as a benchmark for evaluating segmentation performance has inherent limitations. Age is an indirect correlate of PVS burden, and observed associations cannot definitively confirm segmentation accuracy. For example, other age-related pathologies misclassified as PVS, such as lacunes, could inflate correlations with age. In addition, global age-related changes, such as diffuse atrophy or shifts in tissue contrast, might spuriously strengthen age-related correlations. Nevertheless, no evidence of this was found in the test_6_ dataset, where manual removal of false-positive non-PVS voxels had minimal impact on voxel-level precision and no appreciable impact on the strength of age-related associations with PVS burden.

Forth, while lacunes were a relatively uncommon occurrence in the datasets included in this work, their potential misclassification as PVS remains a concern for segcsvd and other automated PVS segmentation tools. Addressing this issue may require additional post-processing steps, such as targeted manual correction, or automated lacune segmentation, which will be considered in future work.

Finally, the WMH mask is implemented as an optional input to segcsvd, incorporating anatomical priors to improve accuracy in individuals with white matter pathology. Ablation-based analyses indicated that excluding the WMH mask had minimal impact on basal ganglia PVS segmentation, a region of strong clinical interest for PVS. Outside the basal ganglia, exclusion resulted in a moderate (~10%) reduction in accuracy, although the magnitude of this effect likely varies by population. In younger or healthier individuals with minimal white matter disease, the model may be robust to the omission of the WMH mask; however, this requires further validation.

## Conclusion

In conclusion, segcsvd provides fully automated segmentation of PVS on routinely acquired T1-weighted images for multisite, diverse datasets. Trained on a large dataset incorporating anatomical priors and extensive augmentation, the model demonstrated strong overall performance and yielded PVS burden estimates that reliably captured expected age associations across diverse cohorts. Robustness tests further demonstrated consistent performance on out-of-distribution data under simulated contrast and noise variation, highlighting advantages over existing benchmark methods.

## Author contributions

1) Conceptualization, 2) data curation, 3) formal analysis, 4) funding acquisition, 5) investigation, 6) methodology, 7) project administration, 8) resources & data acquisition, 9) software, 10) supervision, 11) validation, 12) visualization, 13) writing – original draft, 14) writing – review and editing

EG: 1, 3, 6, 9, 12, 13, 14

JR: 1, 2, 5, 6, 7, 10, 14

LAW: 2, 11, 12, 14

SB: 2, 7, 11, 14

JO: 1, 11, 14

CJMS: 2, 7, 8, 14

VY: 2, 7, 14

FG: 2, 10, 11, 14

RDC: 1, 2, 4, 8, 14

MVH: 1, 2, 4, 8, 14

AEL: 1, 4, 5, 7, 8, 14

CMT: 1, 4, 5, 7, 8, 14

SK: 1, 4, 5, 7, 8, 14

MAB: 1, 2, 4, 5, 7, 8, 14

RB: 1, 2, 4, 5, 7, 8, 10, 14

SS: 1, 2, 4, 8, 14

RHS: 1, 4, 5, 7, 8, 10, 14

MM: 1, 4, 5, 7, 8, 14

NS: 1, 2, 4, 5, 7, 8, 14

AM: 1, 2, 4, 5, 7, 8, 10, 14

BJM: 1, 2, 4, 5, 7, 8, 10, 14

JMW: 1, 4, 5, 8, 10, 14

SEB: 1, 4, 5, 8, 10, 14

ASPL: 1, 4, 5, 8, 10, 14

MG: 1, 4, 5, 8, 10, 14

### Acknowledgements

We thank all the study participants for their time and valuable contribution to our studies, which would not be possible without their support. This work was supported by Canadian Institutes of Health Research PJT173387 and PJT190220. We acknowledge the Weston-Selfridges UK Brain Institute and Fondation Leducq Transatlantic Network of Excellence for their support on PVS-related studies. We thank the support from the Dr. Sandra E. Black Centre for Brain Resilience and Recovery. **MG is supported by the Canada Research Chairs program (CRC-2021-00374). JMW, MH, and RDC acknowledge the UK Dementia Research Institute which is funded by the UK Medical Research Council, Alzheimer’s Society and Alzheimer’s Research UK, and the Row Fogo Centre for Research into Ageing and the Brain, funded by the Row Fogo Charitable Trust. SK acknowledges research support from Brain and Behavior Foundation, National institute on Ageing, BrightFocus Foundation, Brain Canada, Canadian Institute of Health Research, Canadian Consortium on Neurodegeneration in Aging, Centre for Ageing and Brain Health Innovation, Centre for Addiction and Mental Health, and an Academic Scholars Award from the Department of Psychiatry, University of Toronto. Equipment support from Soterix Medical. JO acknowledges funding by the Alzheimer’s Association (24AARF-1242638) and BrightFocus (A2024012F)**.

## Data and code availability statement

A portion of the data used in this study is available through formal data request processes (ADNI, CAHHM). Data from the remaining three studies (LD, CAIN, ONDRI) was collected under institutional ethics approvals that do not permit public sharing. The code, models, and detailed usage instructions for segcsvd are openly available on GitHub, accessible via the following link: https://github.com/AICONSlab/segcsvd.

## References

Ballerini L, Lovreglio R, Valdés Hernández MDC, Ramirez J, MacIntosh BJ, Black SE, Wardlaw JM. 2018. Perivascular Spaces Segmentation in Brain MRI Using Optimal 3D Filtering. Scientific Reports. 8:2132. doi: 10.1038/s41598-018-19781-5.

Bernal J, Valdés-Hernández MDC, Escudero J, Duarte R, Ballerini L, Bastin ME, Deary IJ, Thrippleton MJ, Touyz RM, Wardlaw JM. 2022. Assessment of perivascular space filtering methods using a three-dimensional computational model. Magnetic Resonance Imaging, 93: 33–51. 10.1016/j.mri.2022.07.016.

Billot B, Magdamo C, Cheng Y, Arnold SE, Das S, Iglesias JE. 2023. Robust machine learning segmentation for large-scale analysis of heterogeneous clinical brain MRI datasets. Proceedings of the National Academy of Sciences 120:e2216399120. doi:10.1073/pnas.2216399120.

Boone L, Biparva M, Mojiri Forooshani P, Ramirez J, Masellis M, Bartha R, Symons S, Strother S, Black SE, Heyn C, Martel AL, Swartz RH, Goubran M. 2023. ROOD-MRI: Benchmarking the robustness of deep learning segmentation models to out-of-distribution and corrupted data in MRI. Neuroimage 278:120289. doi:10.1016/j.neuroimage.2023.120289.

Boutinaud P, Tsuchida A, Laurent A, Adonias F, Hanifehlou Z, Nozais V, Verrecchia V, Lampe L, Zhang J, Zhu YC, Tzourio C, Mazoyer B, Joliot M. 2021. 3D segmentation of perivascular spaces on T1-weighted 3 Tesla MR images with a convolutional autoencoder and a U-shaped neural network. Frontiers in Neuroinformatics, 15:641600. doi: 10.3389/fninf.2021.641600. PMID: 34262443; PMCID: PMC8273917.

Choi Y, Nam Y, Choi Y, Kim J, Jang J, Ahn KJ, Kim BS, Shin NY. 2020. MRI-visible dilated perivascular spaces in healthy young adults: A twin heritability study. Human Brain Mapping, 41(18):5313–5324. doi: 10.1002/hbm.25194.

Duarte Coello R, Valdés Hernández MC, Zwanenburg JJM, van der Velden M, Kuijf HJ, De Luca A, Moyano JB, Ballerini L, Chappell FM, Brown R, Biessels GJ, Wardlaw JM. 2024. Detectability and accuracy of computational measurements of in-silico and physical representations of enlarged perivascular spaces from magnetic resonance images. Journal of Neuroscience Methods, 403:110039. doi: 10.1016/j.jneumeth.2023.110039.

Fischl B, Salat DH, van der Kouwe AJ, Makris N, Ségonne F, Quinn BT, Dale AM. 2002. Whole brain segmentation: automated labeling of neuroanatomical structures in the human brain. Neuron 33:341–355.

Frangi AF, Niessen WJ, Vincken KL, Viergever MA. 1998. Multiscale vessel enhancement filtering. Medical Image Computing and Computer-Assisted Intervention — MICCAI’98 Lecture Notes in Computer Science 1496:130.

Gibson E, Ramirez J, Woods LA, Ottoy J, Berberian S, Scott CJM, Yhap V, Gao F, Coello RD, Valdes Hernandez M, Lang AE, Tartaglia CM, Kumar S, Binns MA, Bartha R, Symons S, Swartz RH, Masellis M, Singh N, Moody A, MacIntosh BJ, Wardlaw JM, Black SE, Lim ASP, Goubran M; ONDRI Investigators, ADNI, CAIN Investigators, colleagues from the Foundation Leducq Transatlantic Network of Excellence. 2024. segcsvdWMH: A Convolutional Neural Network-Based Tool for Quantifying White Matter Hyperintensities in Heterogeneous Patient Cohorts. Human Brain Mapping. 45(18):e70104. doi: 10.1002/hbm.70104.

Huang P, Liu L, Zhang Y, Zhong S, Liu P, Hong H, Wang S, Xie L, Lin M, Jiaerken Y, Luo X, Li K, Zeng Q, Cui L, Li J, Chen Y, Zhang R; Alzheimer’s Disease Neuroimaging Initiative. 2024. Development and validation of a perivascular space segmentation method in multi-center datasets. Neuroimage 298, 1053–8119.

Isensee F, Jaeger PF, Kohl SAA, Petersen J, Maier-Hein KH. 2021. nnU-Net: a self-configuring method for deep learning-based biomedical image segmentation. Nature Methods 18(2):203–211. doi: 10.1038/s41592-020-01008-z. Epub 2020 Dec 7. PMID: 33288961.

Merveille O, Naegel B, Talbot H, Najman L, Passat N. 2017. 2D filtering of curvilinear structures by ranking the orientation responses of path operators (RORPO). Image Processing On Line 7:246–261. doi:10.5201/ipol.2017.207.

Ntiri EE, Holmes MF, Forooshani PM, Ramirez J, Gao F, Ozzoude M, Adamo S, Scott CJM, Dowlatshahi D, Lawrence-Dewar JM, Kwan D, Lang AE, Symons S, Bartha R, Strother S, Tardif JC, Masellis M, Swartz RH, Moody A, Black SE, Goubran M. 2021. Improved Segmentation of the Intracranial and Ventricular Volumes in Populations with Cerebrovascular Lesions and Atrophy Using 3D CNNs. Neuroinformatics, 19(4):597–618. doi: 10.1007/s12021-021-09510-1.

Pham W, Lynch M, Spitz G, O’Brien T, Vivash L, Sinclair B, Law M. 2022. A critical guide to the automated quantification of perivascular spaces in magnetic resonance imaging. Frontiers in Neuroscience 16:1021311. doi:10.3389/fnins.2022.1021311. PMID: 36590285; PMCID: PMC9795229.

Maier-Hein L, Reinke A, Godau P, Tizabi MD, Buettner F, Christodoulou E, Glocker B, Isensee F, Kleesiek J, Kozubek M, Reyes M, Riegler MA, Wiesenfarth M, Kavur AE, Sudre CH, Baumgartner M, Eisenmann M, Heckmann-Nötzel D, Rädsch T, Acion L, Antonelli M, Arbel T, Bakas S, Benis A, Blaschko MB, Cardoso MJ, Cheplygina V, Cimini BA, Collins GS, Farahani K, Ferrer L, Galdran A, van Ginneken B, Haase R, Hashimoto DA, Hoffman MM, Huisman M, Jannin P, Kahn CE, Kainmueller D, Kainz B, Karargyris A, Karthikesalingam A, Kofler F, Kopp-Schneider A, Kreshuk A, Kurc T, Landman BA, Litjens G, Madani A, Maier-Hein K, Martel AL, Mattson P, Meijering E, Menze B, Moons KGM, Müller H, Nichyporuk B, Nickel F, Petersen J, Rajpoot N, Rieke N, Saez-Rodriguez J, Sánchez CI, Shetty S, van Smeden M, Summers RM, Taha AA, Tiulpin A, Tsaftaris SA, Van Calster B, Varoquaux G, Jäger PF. 2024. Metrics reloaded: recommendations for image analysis validation. Nature Methods 21:195–212. doi:10.1038/s41592-023-02151-z.

McDonnell MD, Abbott D. 2009. What is stochastic resonance? Definitions, misconceptions, debates, and its relevance to biology. PLoS Computational Biology 5(5):e1000348. doi:10.1371/journal.pcbi.1000348. PMID: 19562010; PMCID: PMC2660436.

Mojiri P, Biparva M, Ntiri EE, Ramirez J, Boone L, Holmes M, Adamo S, Gao F, Ozzoude M, Scott C, Dowlatshahi D, Lawrence-Dewar J, Kwan D, Lang A, Marcotte K, Leonard C, Rochon E, Heyn C, Bartha R, Strother S, Tardif JC, Symons S, Masellis M, Swartz R, Moody A, Black SE, Goubran M. 2022. Deep Bayesian networks for uncertainty estimation and adversarial resistance of white matter hyperintensity segmentation. Human Brain Mapping, 43(7):2089–2108. doi: 10.1002/hbm.25784

Perona P, Malik J. 1990. Scale space and edge detection using anisotropic diffusion. IEEE Transactions on Pattern Analysis and Machine Intelligence 12:629–639.

Sinclair B, Vivash L, Moses J, Lynch M, Pham W, Dorfman K, Marotta C, Koh S, Bunyamin J, Rowsthorn E, Jarema A, Peiris H, Chen Z, Shultz SR, Wright DK, Kong D, Naismith SL, O’Brien TJ, Law M. 2024. Perivascular space Identification Nnunet for Generalised Usage (PINGU). arXiv preprint. Available from: https://arxiv.org/abs/2405.08337.

Wardlaw JM, Smith C, Dichgans M. 2019. Small vessel disease: mechanisms and clinical implications. The Lancet Neurology 18:684–696. doi:10.1016/S1474-4422(19)30079-1.

Wardlaw JM, Benveniste H, Nedergaard M, Zlokovic BV, Mestre H, Lee H, Doubal FN, Brown R, Ramirez J, MacIntosh BJ, Tannenbaum A, Ballerini L, Rungta RL, Boido D, Sweeney M, Montagne A, Charpak S, Joutel A, Smith KJ, Black SE; colleagues from the Fondation Leducq Transatlantic Network of Excellence on the Role of the Perivascular Space in Cerebral Small Vessel Disease. 2020. Perivascular spaces in the brain: anatomy, physiology and pathology. Nature Reviews Neurology, 16(3):137–153. doi: 10.1038/s41582-020-0312-z.

Wardlaw JM, Smith EE, Biessels GJ, Cordonnier C, Fazekas F, Frayne R, Lindley RI, O’Brien JT, Barkhof F, Benavente OR, Black SE, Brayne C, Breteler M, Chabriat H, Decarli C, de Leeuw FE, Doubal F, Duering M, Fox NC, Greenberg S, Hachinski V, Kilimann I, Mok V, van Oostenbrugge R, Pantoni L, Speck O, Stephan BCM, Teipel S, Viswanathan A, Werring D, Chen C, Smith C, van Buchem M, Norrving B, Gorelick PB, Dichgans M. 2013. STandards for ReportIng Vascular changes on nEuroimaging (STRIVE v1). The Lancet Neurology, 12:822–838.

Waymont JMJ, Valdés Hernández MDC, Bernal J, Duarte Coello R, Brown R, Chappell FM, Ballerini L, Wardlaw JM. Systematic review and meta-analysis of automated methods for quantifying enlarged perivascular spaces in the brain. 2024. Neuroimage. 297:120685. doi: 10.1016/j.neuroimage.2024.120685.

